# BC-Predict: Mining of signal biomarkers and multilevel validation of cascade classifier for early-stage breast cancer subtyping and prognosis

**DOI:** 10.1101/2024.03.20.24304604

**Authors:** Sangeetha Muthamilselvan, Ashok Palaniappan

## Abstract

Disease heterogeneity is the hallmark of breast cancer, which remains a scourge and the most common malignancy among women. With a steep increase in breast cancer morbidity and mortality, there exists a critical need for effective early-stage theragnostic and prognostic biomarkers. This would help in patient stratification and optimal treatment selection towards better disease management. In this study, we examined four key problems with respect to the characterization of breast cancer heterogeneity, namely: (i) cancer screening; (ii) identification of metastatic cancers; (iii) molecular subtype (TNBC, HER2, or luminal); and (iv) histological subtype (ductal or lobular). We mined the available public-domain transcriptomic data of breast cancer patients from the TCGA and other databases using stage-encoded statistical models of gene expression, and identified stage-salient, monotonically expressed, and problem-specific biomarkers. Next we trained different classes of machine learning algorithms targeted at the above problems and embedded in these feature spaces. Hyperparameters specific to each algorithm were optimized using 10-fold cross-validation on the training dataset. The optimized models were evaluated on the holdout testset to identify the overall best model for each problem. The best model for each problem was validated with: (i) multi-omics data from the same cohort (miRNA and methylation profiles); (ii) external datasets from out-of-domain cohorts; and (iii) state of the art, including commercially available breast cancer panels. External validation of our models matched or bested available benchmarks in the respective problem domains (balanced accuracies of 97.42% for cancer vs normal; 88.22% for metastatic v/s non metastatic; 88.79% for ternary molecular subtyping; and ensemble accuracy of 94.23% for histological subtyping). We have translated the results into BC-Predict, a freely available web-server that forks the best models developed for each problem, and provides the cascade annotation of input instance(s) of expression data, along with uncertainty estimates. BC-Predict is meant for academic use and has been deployed at: https://apalania.shinyapps.io/BC-Predict.

## Introduction

Breast cancer is the most common cancer in women, accounting for 32% of all female cancers globally and 28.2% of female cancers in India^1^. With about 2.3 million new cases globally in 2020 (11.7% of total), its incidence surpasses that of lung cancer. The statistics paint a grim portrait of burden of disease: 1 in 4 cancer cases and 1 in 6 cancer deaths globally could be attributed to breast cancer, with 88% higher incidence in transitioned countries relative to transitioning countries^2^. The risk of a person developing breast cancer depends on many factors like sex (women account for >99.5%), age (>80% occur in postmenopausal women), high-risk family history (upto 30% of cases), and genetic factors. The interplay between weak susceptibility alleles and the other risk factors is key to the etiology of the ‘cancer phenotype’^3,4^. Genetic loci with predisposing mutations include: BRCA1/ BRCA2 (autosomal dominant, 50-85% life time risk)^5^, TP53 (Li-Fraumeni syndrome, 80-90% life time risk)^6^, CDH1 (60% life time risk and primarily lobular subtype), STK11 (Peutz-Jeghers syndrome, 50% risk), PTEN (Cowden syndrome with 20-50% risk^7^; Lynch syndrome with 25% risk), PALB2 (partner and localiser to BRCA2, age-dependent risk), ATM, BRIP1, CHEK2 (all about 20% risk) and RAD51C/RAD51D (14-20% risk). The modifiable lifestyle risk factors include physical inactivity especially post-menopausal obesity (100% additional risk), smoking (24% more risk), alcohol (7% risk for every 10g/day), and combined Hormone Replacement therapy (∼20% further risk depending on length of use/stop)^8^. The prevalence of the risk factors varies by country and region. The typical onset of breast cancer is 60-70 years in western countries, but appears to be anticipated at 40-50 years in countries like India^9^. Data maintained at national registries suggest that the urbanization and growth of cities, ‘modernized’ food habits (e.g, high consumption of ultra-processed foods), and lifestyle changes have contributed to the increased incidence of breast cancer in urban areas, whereas betel quid and tobacco chewing habits have significantly contributed to its incidence in rural areas (*P* = 0.003)^10^. These cancers tend to be more aggressive with poorer prognosis (higher grade/size, lymphovascular-invasion positive, triple negative, HER2 positive, node positive, and medullary/metaplastic/micro-papillary/pleomorphic sub-types). The frequent presentation of breast cancer in its advanced and less treatable stages in traditional societies could be traced partly to the inadequate social awareness and extant taboos, leading to subpar survival outcomes. Such conditions compound gender inequalities, stereotypes and the burden of disease for whole families, and call for remediation of the situation.

Due to the complexity associated with cancers, a composite feature space is necessary to capture the transformation of cells and subsequent disease progression. This may be balanced with the curse of dimensionality that dominates machine learning. AI models based on whole-genome or whole-exome sequencing may be impractical and uninterpretable. McKinney et al. have developed a mammogram-based AI model for breast cancer screening rivalling radiologist readings, paving the way for AI-based decision support systems^11^. Convolutional neural network (CNN) models have been developed for identifying breast cancer samples as well as cancer subtyping based on 7091 genes^12^. CUP-AI-DX includes two models: 1D inception CNN model for classifying cancers of unknown primary based on 817 expression features; and (ii) Random Forest model for breast cancer subtyping based on 5925 expression features^13^. Breast cancer subtyping models include learning on PAM50 inferred labels^14^ via either functional spectra of gene expression profiles^15^ or deep convolution of RNAseq and CNV profiles^16^. Significant strides have been made towards mechanistic understanding and treatment of breast cancer, which has the most number of FDA-approved molecular panels aimed at early-stage actionable information about the disease. These biomarker panels include OncotypeDx based on TAILORx and RxPONDER studies^17^, EndoPredict and EndoPredict Plus^18^, MammaPrint^19^, Prosigna (based on PAM50 and OPTIMA study)^20^, and Breast Cancer Index^21^. Decision aids like PREDICT, Nottingham Prognostic Index (NPI) and Adjuvant Online based on IHC4 (ER/PR/HER2/Ki67) or IHC4+C (including clinical/pathological features like age, tumour size, grade and nodal status) parameters define the level of clinical risk for adjuvant chemotherapy without relying on tumour profiling tests. The translation of AI models into software-as-medical-devices holds promise for bridging health disparities^22^.

The heterogeneity of breast cancer poses formidable challenges, and individual cancer manifestations vary so much that the available biomarker panels retain validity only in limited settings, thereby leaving a large cohort indeterminate^23^. Changes in gene expression and mutations modifying protein activities are etiological molecular events driving the cancer phenotype^24^. An integrated precision-medicine approach to early detection, effective therapy and favourable prognosis is necessary. Techniques from the field of machine learning could be highly effective in discerning key features in complex datasets, including gene expression datasets, and learning models that map these features to crucial clinical outcomes related to the diagnosis, prognosis, and treatment of cancers^25^. Unsupervised learning techniques have been used to identify subtypes in breast cancer based on gene expression^26^. The molecular subtype of breast cancer could influence the choice of adjuvant therapy^27,28^. Among the histological subtypes, invasive lobular carcinoma is considered indolent and might necessitate a switch to a more lethal treatment regime^29^. Here we have developed a novel framework for identifying the markers of changes in gene expression profiles across the stages and subtypes of breast cancer, enabling means for differential diagnosis and personalized medicine. These candidate biomarkers were applied to create models that address the following challenges in breast cancer heterogeneity: (i) cancer or normal screening; (ii) non-metastatic or metastatic discrimination; (iii) molecular subtyping; and (iv) histological subtyping. Together these models could also enable the prognosis of breast cancer^30,31^. The optimal models for each problem required only a handful of features that can be quantified using qRT-PCR. All the models were integrated into BC-Predict, a web-based unified interface for harnessing the models. BC-Predict is available for academic research at: https://apalania.shinyapps.io/BC-Predict. All the Supplementary Information for this study are available at: https://doi.org/10.6084/m9.figshare.25282906.

## Methods

Problems related to the characterization of breast cancer heterogeneity:

Four problems related to the delineation of individual breast cancers with respect to the expression data of patient samples were considered:

1. Is the patient sample ‘cancer’ or ‘normal’?
2. If cancer: predict ‘non-metastatic’ (stages I, II or III) or ‘metastatic’ (stage-IV cancer).
3. If cancer: predict the molecular subtype of the cancer.
4. If cancer: predict the histological subtype of the cancer.

A generalized workflow for the problems is depicted in Fig. 1.

**Figure 1.**
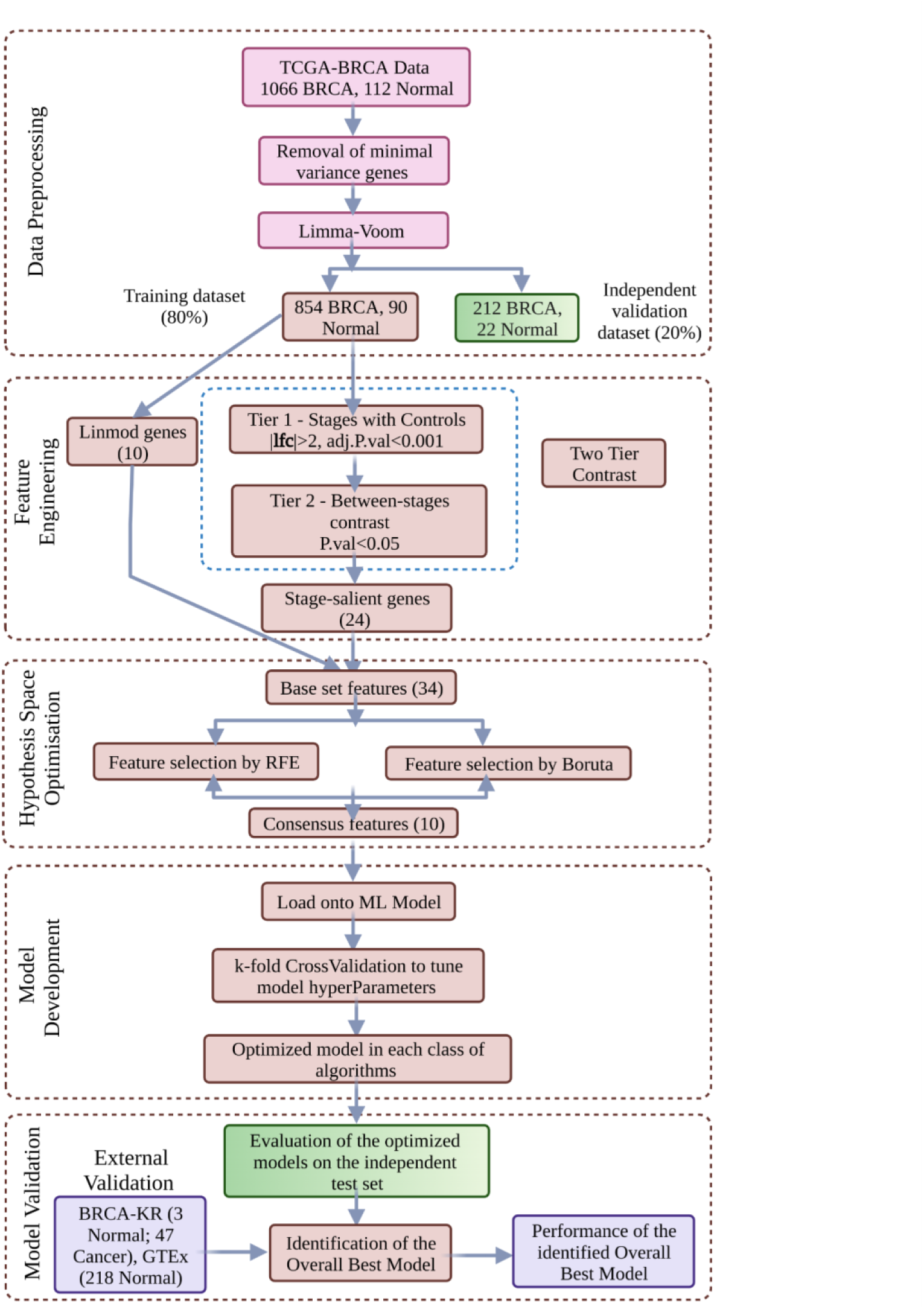
ML model development for Cancer vs. Normal binary classification. Data-driven optimization of a multi-phase workflow is shown. Problem-specific changes to the workflow yield adaptations to the other problems considered in this study.

### Dataset preprocessing

Preprocessing was done in a manner similar to Sarathi and Palaniappan^32^. The source dataset for all problems modeled here was obtained from the TCGA. Normalised BRCA expression data was acquired using the firebrowse portal^33^ (gdac.broadinstitute.org_BRCA.Merge_rnaseqv2_illuminahiseq_rnaseqv2_unc_edu_Level_3_RSEM_genes_normalized_data.Level_3.2016012800.0.0.tar.gz), and RSEM counts were obtained. The patient barcode was matched with the clinical data (gdac.broadinstitute.org_BRCA.Merge_Clinical.Level_1.2016012800.0.0.tar) to extract the patient.stage_event.pathologic_stage variable values that encode the AJCC TNM staging^34^. The sub-stages were then merged to obtain the macro stage categories. Table 1 shows the distribution of sample stages for the breast cancer samples according to the AJCC staging system. It is noted that early-stage BC indicates TNM stage-I or stage-II cancer. Stage-III BC (including T3N1, T4, N2-3) represents loco-regionally advanced BC, whereas T3N0 represents a borderline diagnosis between stages II and III. For the purposes of our study, stages I, II, and III were combined into the ‘non-metastatic’ class.

**Table 1:**
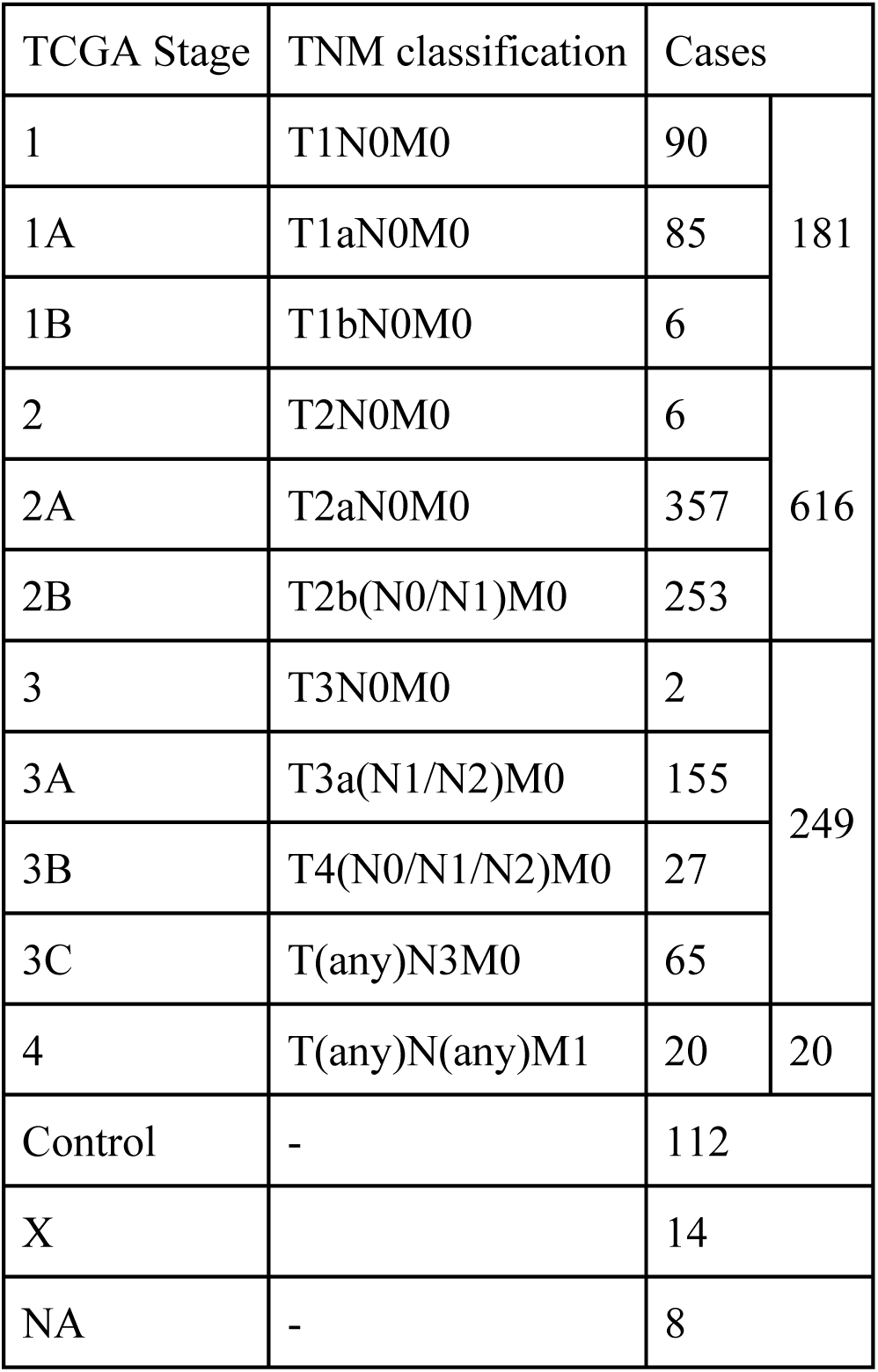
Stage-wise distribution of breast cancer samples in the TCGA data, based on the AJCC system, 2018 revision.

The immunohistochemical (IHC) status of oestrogen receptor (ER) and progesterone receptor (PgR), human epidermal growth factor receptor 2 (HER2) oncogene, and Ki-67 (a marker of cell proliferation) are used together to subtype breast tumors into Triple-negative breast cancer (TNBC), HER2-positive, Luminal A and Luminal B^34,35^, as shown in Table 2. Where reliable Ki-67 measurements are not available, an alternative assessment of tumor proliferation such as tumor grade could be used to distinguish between ‘Luminal A’ and ‘Luminal B’ (which tends to be HER2 negative). Complete ER, PgR and HER2 IHC metadata were available for 719 samples of the TCGA Breast Cancer dataset, and of these, no sample had information on the Ki-67 labeling index nor on the tumor grade, precluding precise differentiation of luminal subtypes of breast cancers into ‘Luminal A’ or ‘Luminal B’. The luminal subtypes A and B were perforce lumped into one ‘Luminal’ type. The 719 samples were accordingly annotated as 567 ‘Luminal’ (generally Luminal A with Grade 1 or 2 and Luminal B with G3), 115 TNBC (generally Grade 3), and 37 HER2 (generally Grade 3) based on the status of ER, PgR and HER2 extracted from the clinical file (Table 2).

**Table 2:**
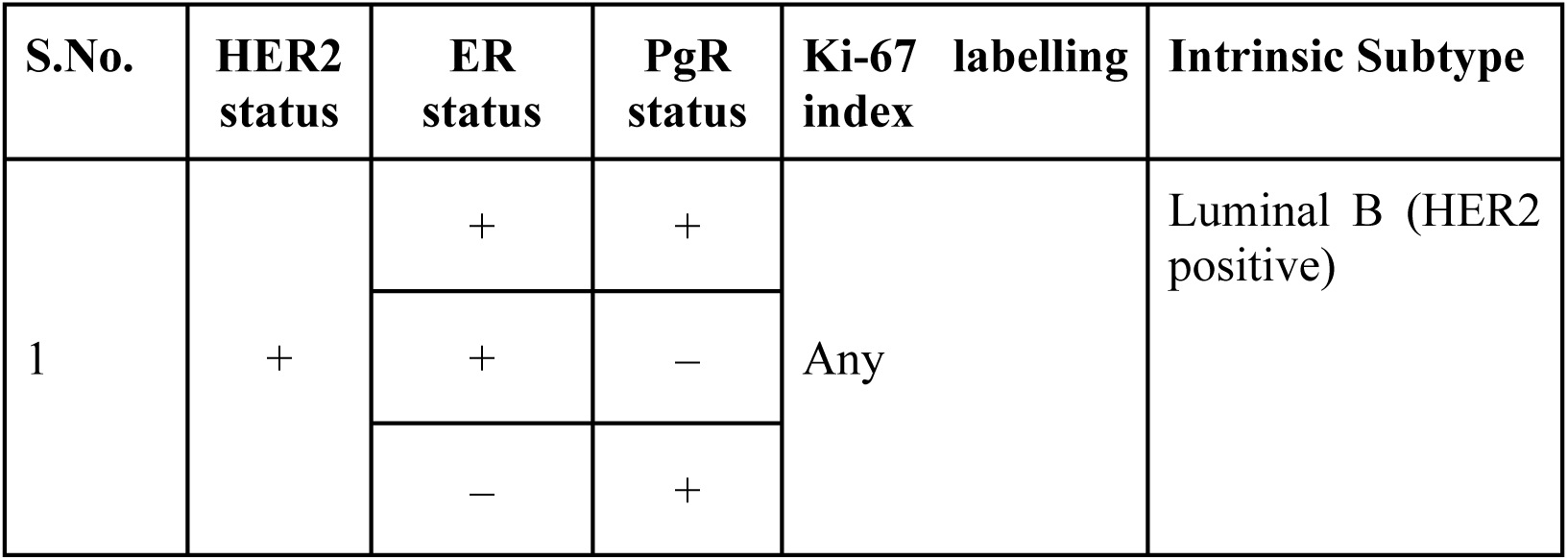
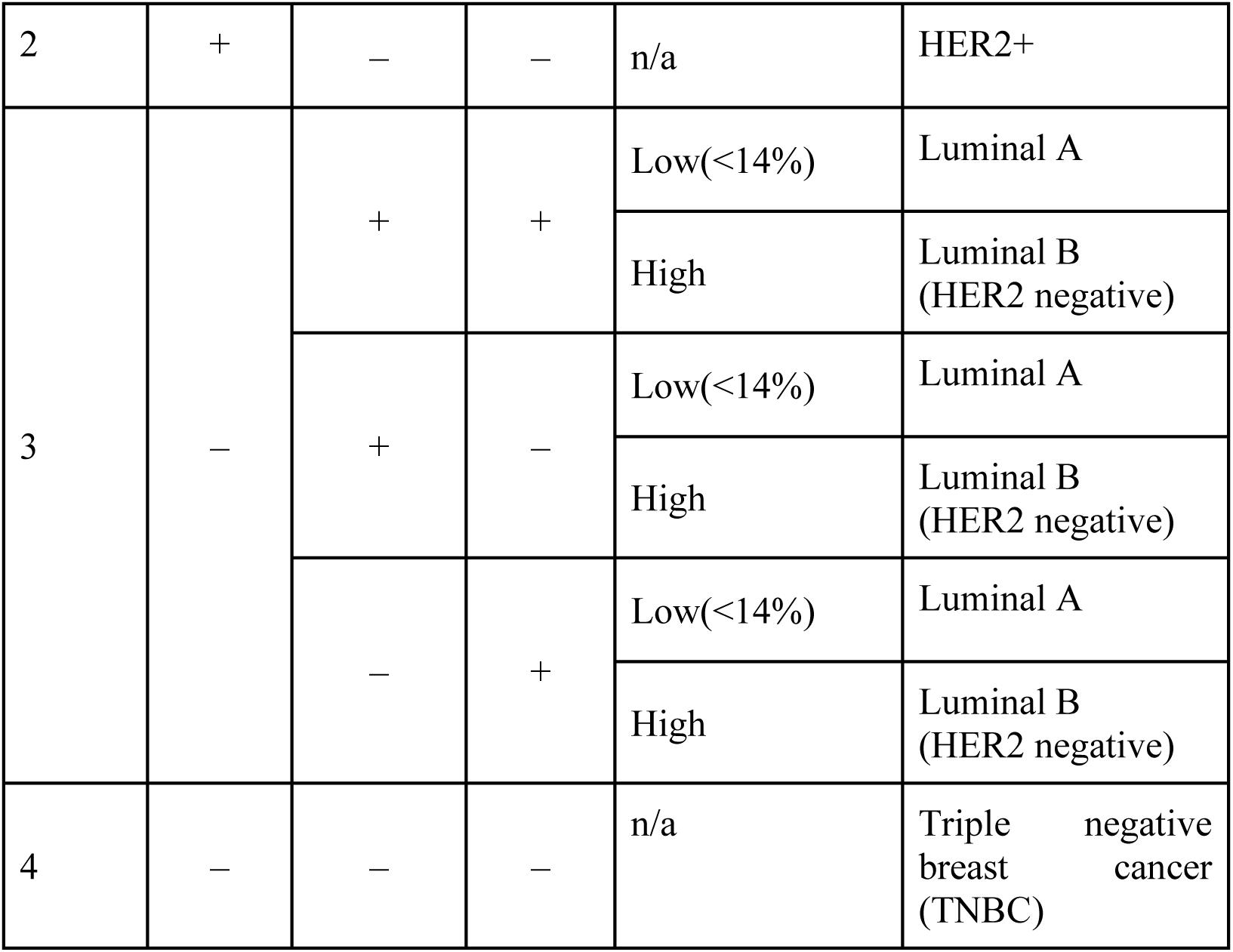
Molecular taxonomy of breast cancer. Luminal A is HER2 negative, whereas Luminal B could be either HER2 positive (accounting for 30% of HER2 positive) or HER2 negative (majority of Luminal B).

| S.No. | HER2 status | ER status | PgR status | Ki-67 labelling index | Intrinsic Subtype |
| --- | --- | --- | --- | --- | --- |
| 1 | + | + | + | Any | Luminal B (HER2 positive) |
|  |  | + | — |  |  |
|  |  | — | + |  |  |
| 2 | + | – | – | n/a | HER2+ |
| 3 | – | + | + | Low(<14%) | Luminal A |
|  |  |  |  | High | Luminal B<br>(HER2 negative) |
|  |  | + | – | Low(<14%) | Luminal A |
|  |  |  |  | High | Luminal B<br>(HER2 negative) |
|  |  | – | + | Low(<14%) | Luminal A |
|  |  |  |  | High | Luminal B<br>(HER2 negative) |
| 4 | – | – | – | n/a | Triple negative<br>breast cancer<br>(TNBC) |

The two most common histological subtypes of breast cancer are infiltrating ductal carcinoma (IDC – no special type) and infiltrating lobular carcinoma (ILC)^36^. ILC tends to be difficult to diagnose, with MR imaging required for determining size and multifocality including contralateral breast (mirror image), and preferential spread to gastrointestinal tract and peritoneum^37^. The sample histological subtype is encoded in the clinical metadata ‘patient.histological_type’ with the major values being, ‘infiltrating ductal carcinoma (IDC)’ and ‘infiltrating lobular carcinoma (ILC)’, and minor values including ‘mixed histology’, ‘metaplastic carcinoma’, ‘mucinous carcinoma’, ‘medullary carcinoma’, and ‘other (specify)’.

Genes that had minimal variation in expression across the samples (i.e, σ < 1) were removed. Cancer samples which were missing stage annotation details were removed. The expression dataset was subjected to variance-stabilization using `voom` function in `limma`^38^. Linear modeling was then performed. The resulting dataset was split 80:20 into a training set and a holdout testset stratified on the outcome variable of each problem. It is noted that the training dataset for Problem #2 suffered an imbalance in the distribution of the outcome classes (16 metastatic vs 837 non-metastatic samples)., which prompted the application of SMOTE correction^39^ (Synthetic Minority Oversampling TEchnique; with arguments: perc.over-represented = 1000 % and perc.under-represented = 300 %). Data preprocessing and analysis was done using R (www.r-project.org). The annotated pre-processed final dataset is available as Supplementary File S1.

### Construction of feature space

Feature spaces for each problem were constructed using only the training dataset. Initially the differential expression of genes across cancer stages relative to healthy samples was studied using linear modelling with the limma package^40^. A two-leveled contrast protocol, viz. level-I: stage vs control and level-II: inter-stages contrast,^22^ was then applied to produce the following classes of biomarkers:

(1) Stage-salient genes, obtained from all possible pairwise contrasts between the cancer stages; and
(2) Monotonically expressed genes, obtained from strictly increasing or strictly decreasing mean expression across the cancer stages.

In addition, expression contrasts specific to the problem under consideration were used, namely:

(1) contrast of non-metastatic vs metastatic cancers;
(2) three-way pairwise contrasts between the molecular subtypes; viz. (i) Luminal vs HER2, (ii) Luminal vs TNBC & (iii) HER2 vs TNBC;
(3) contrast of ductal vs lobular histologies.

The above strategies yielded problem-specific chimeric feature spaces that could span the informative dimensions in each case.

### Building problem-specific classification models

A composite feature space comprising the top-ranked genes from the linear model, stage-salient genes, and genes from the problem-specific contrast was subjected to the consensus of two feature selection techniques: (i) Boruta, a wrapper algorithm using Random Forest to select features based on a measure of importance to the outcome variable of interest^41^; and (ii) Recursive Feature Elimination (RFE), a method that uses backward selection passes to trim the space of predictor variables. The workflow of the machine learning model development in Figure 1 presented in the context of cancer v/s normal was adapted for the non-metastatic v/s metastatic and subtype discrimination problems. The training dataset with the final set of features was loaded onto models based on various ML algorithms, including Random forest (ensemble method that builds numerous decision trees and ‘bags’ the majority vote), SVM (geometric method that finds an optimal separating hyperplane in high-dimensional space), k-NN (based on proximal classes), Neural Networks (both 1-layer and 2-layer), and XgBoost (ensemble method that builds a sequence of classifiers boosted on challenging instances). Algorithm-specific hyperparameters were optimized using 10-fold cross-validation on the training dataset. The performance of the hyperparameter-optimized models was evaluated on the holdout testset to identify the overall best model for each problem. Evaluation was performed using a slew of metrics, including balanced accuracy, F1-score, area under ROC (AUROC), Mathews’ correlation coefficient (MCC), and Positive Predictive Value (PPV).

### Validation

The overall best model for each problem was validated primarily by performing inference on out-of-domain external datasets. Table 3 shows the datasets used in the development and validation of the ML models for the respective classification problems. In addition, we sought to obtain concurrence for our models from multi-omic signatures, as discussed below.

(1) External validation:
  (i) Normal vs. cancer We validated model#1 on multiple independent external breast cancer datasets:
    (a) BRCA-KR dataset retrieved from the ICGC DataPortal (https://dcc.icgc.org/) using ‘BRCA’ as the search keyword^42^, containing 47 cancer samples and 3 control samples.
    (b) GTEx normal breast dataset (by querying for ‘Breast’ in the “GTEX_phenotype primarysite”)^43^ with 218 control samples.
    (c) GSE18549, GSE211167, and METABRIC datasets.
  (ii) Non-metastatic vs. metastatic We validated model#2 on two different external breast cancer datasets:
    (a) BRCA-KR dataset described above, with all 47 cancer samples being non-metastatic cancers.
    (b) GSE18549 dataset of metastatic cancers (https://www.ncbi.nlm.nih.gov/geo/query/acc.cgi?acc=GSE18549)^44^, with 14 samples having ‘Breast’ as the primary tumor site.
  (iii) Molecular subtyping We validated model#3 on two different external breast cancer datasets:
    (a) METABRIC a landmark study of breast cancer transcriptomics, available on cBioPortal (https://www.cbioportal.org/study/summary?id=brca_metabric)^45^. Breast cancer samples in METABRIC were subtyped as Luminal, HER2, or TNBC based on the IHC status of ER, PgR and HER2 extracted from the METABRIC clinical metadata. This yielded 1415 Luminal, 127 HER2, and 299 TNBC METABRIC samples. Since METABRIC had used microarray technology to measure gene expression, a platform-specific bias might be induced. To mitigate this bias and obtain data compatible with RNA-Seq technology, we applied the Feature Specific Quantile Normalization (FSQN) technique to the METABRIC data^46^.
    (b) GEO Dataset GSE211167^47^, consisting of only TNBC samples from 26 patients of African ancestry. The dataset was log_2_-transformed prior to serving for model inference.
  (iv) Histological subtyping: We validated model#4 on an external breast cancer dataset from cBioPortal with 96 IDC and 19 ILC samples (https://www.cbioportal.org/study/summary?id=brca_mbcproject_wagle_2017)^48^.
(2) Multi-omics:
  (i) Integration of miRNA analysis miRNAs play a crucial role in the regulation of global mRNA expression in both physiological and pathological processes, including the invasion and metastasis of cancer. By exerting control over the expression of target genes, miRNAs act as oncogenes, tumor-suppressive genes, and modulators of distant metastasis in breast cancer. To identify differentially expressed (DE) miRNAs, we used the miRSeq dataset from the same TCGA BRCA cohort (gdac.broadinstitute.org_BRCA.Merge_mirnaseq illuminahiseq_mirnaseq bcgsc_ca Level_3 miR_isoform_expression data.Level_3.2016012800.0.0.tar.gz). Being a transcriptomics dataset, the miRSeq dataset was treated akin to the mRNASeq dataset, with cancer stage as indicator variable. DE stage-specific miRNAs were revealed upon application of the two-level contrast (stage vs control level-I contrast and inter-stages level-II contrast). For each identified stage-salient miRNA, the target genes were predicted using multiMiR^49^, which provides an integration of 14 miRNA-mRNA interaction databases including TargetScan^50^, miRDB^51^, miRanda^52^, and miRTarBase^53^. Of the predicted targets for each miRNA, the stage-salient targets were investigated for differential miRNA expression-driven genes.
  (ii) Identification of differential methylation-driven genes (DMDGs)

Epigenetic processes such as methylation could contribute to changes in gene expression, and drive pathological processes. To evaluate differentially methylated genes, we used the Level3-processed 450k methylation dataset from the same TCGA BRCA cohort (gdac.broadinstitute.org_BRCA.Merge_methylation humanmethylation450 jhu_usc_edu__Level_3 within_bioassay_data_set_function data.aux.2016012800.0.0.tar.gz). The correlation between methylation and expression of the stage-salient genes was analyzed using R MethylMix^54^, with the preset threshold –0.3 and p-value < 0.001. Differentially methylated states were identified using significance from Wilcoxon rank-sum testing (adj. p.value < 0.05) with an additional effect size filter (> 0.1). Genes passing these marker filters were designated as differential methylation-driven genes. Stage-salient differentially methylated genes were identified using the consensus of three stage-informed models, namely Averep, M-value and MethylMix as described in ref. 55.

**Table 3.**
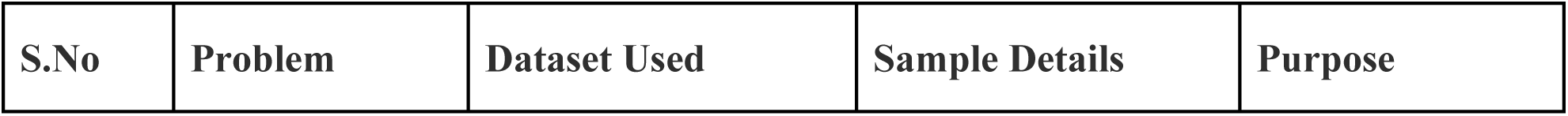
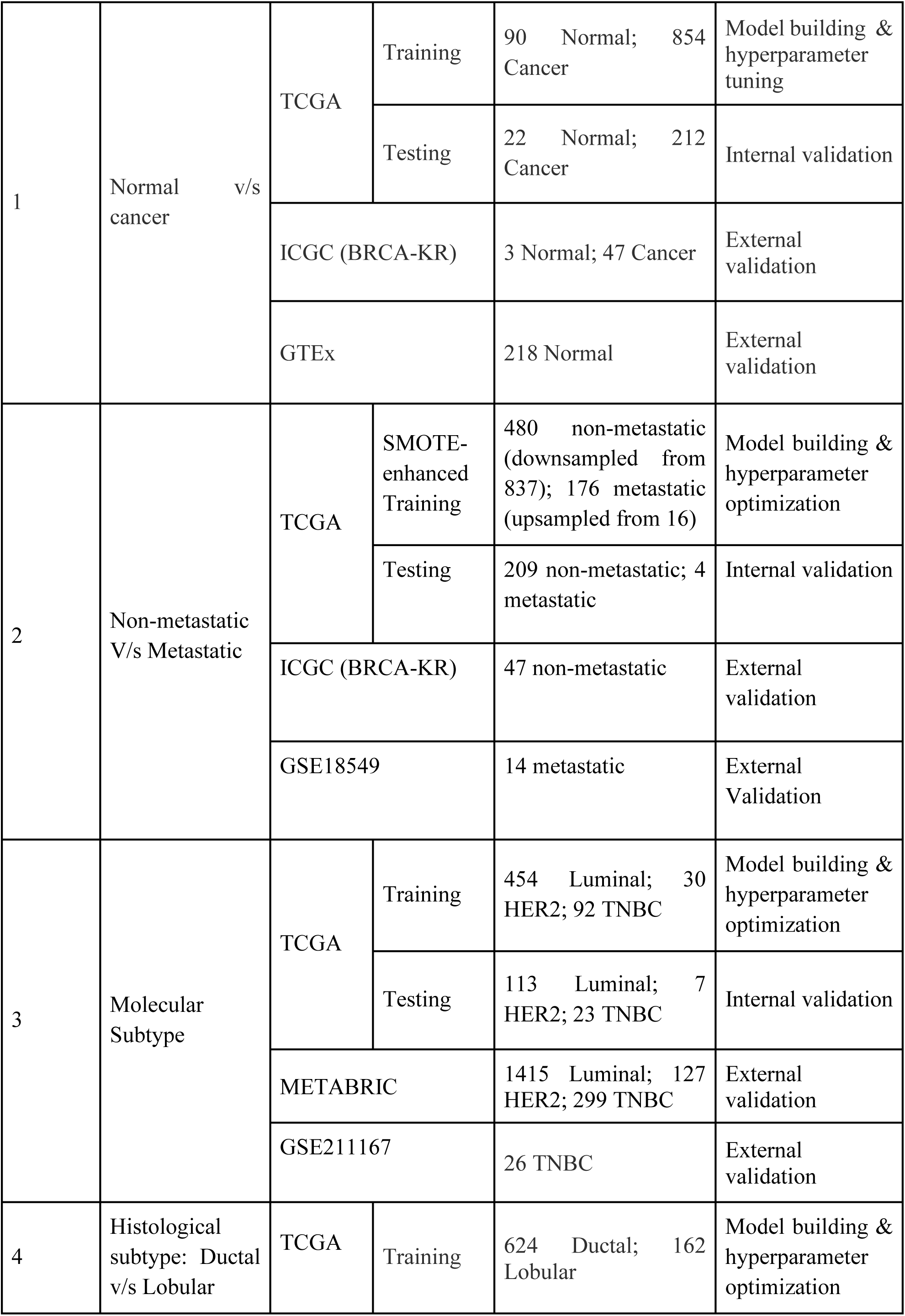
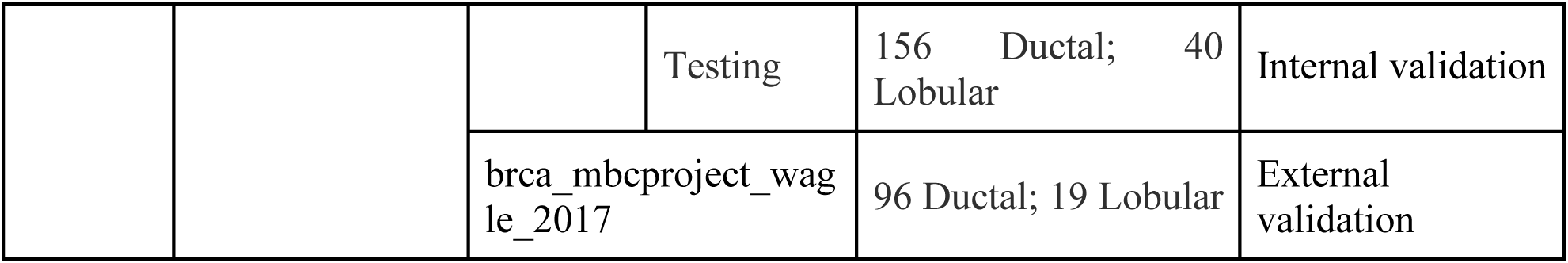
Datasets used in the modelling of BRCA classification problems. The ‘normal vs cancer’ model is additionally validated on GSE18549, GSE211167, and METABRIC datasets.

### Development of cascade classifier

A prediction pipeline that integrates the predictions from all the models into one combined readout was designed. A schematic for one such cascade model is shown in Figure 2. Based on the decision at the shown fork, the new sample may be taken forward for assessment of metastatic potential and molecular / histological subtyping. The final readout for a sample from the cascade classifier would consolidate the inference from each model; for e.g., ‘Metastatic triple-negative ductal cancer’. This formed the basis for the development of BC-Predict.

**Figure 2.**
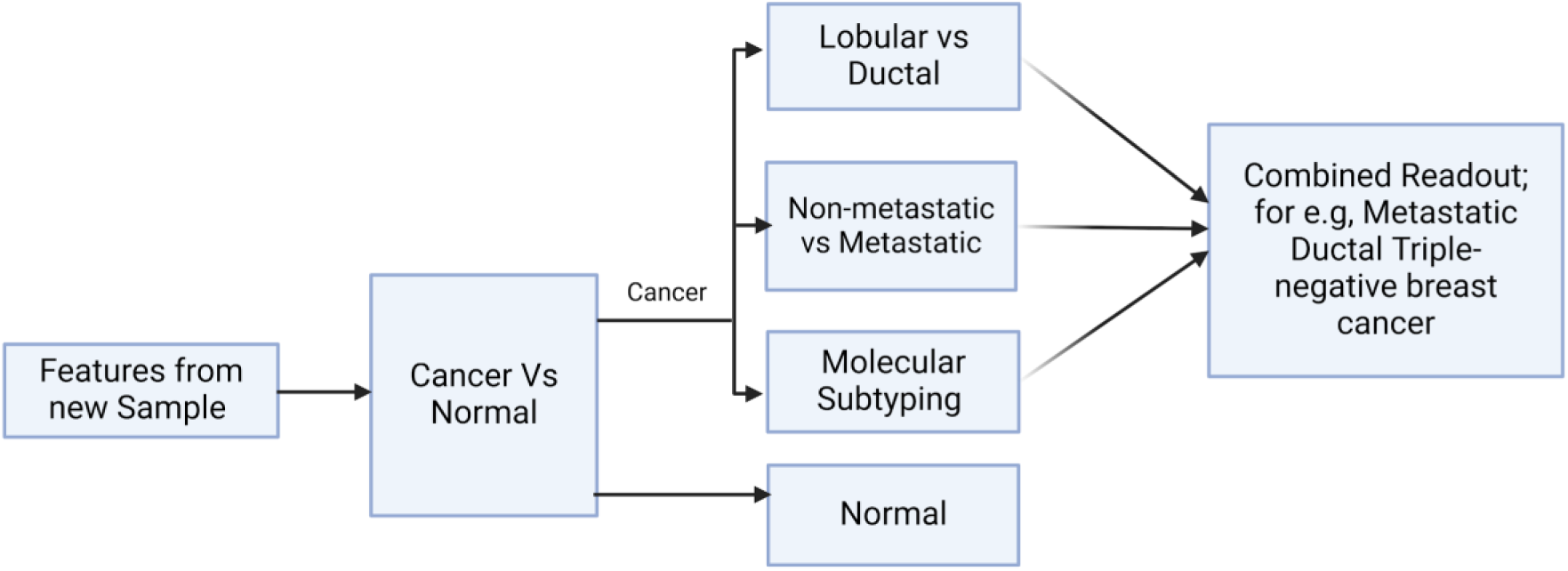
Design of BC-Predict. A schematic of a cascade model for early-stage breast cancer subtyping and prognosis is presented. If the sample is predicted as ‘cancer’ in the first level, it is passed through three more models in the second level that characterize the cancer sample towards personalized prognosis.

## Results

The TCGA BRCA dataset consisted of 1212 samples, each with the measurement of expression of 20532 genes. Post data preprocessing, we obtained an annotated dataset of 1178 samples x 18880 genes (Supplementary File S1). An adj.p.value cut-off of 0.05 yielded 14838 DE genes in breast cancer samples. Tightening the significance to 1E-05 still yielded 10167 DE genes, underscoring the persistence of genome instability in the march of cancer^4^. A volcano plot depicting differentially expressed genes showed significant dispersion (Figure 3a), meaning some genes were much more dysregulated than others. We performed a principal components analysis with the top ten genes from the linear modelling, and found that a clear separation between the normal and cancer samples could be obtained (Figure 3b). This provided some basis for considering top-ranked genes from the linear modeling as candidate cancer-specific biomarkers. Table 4 provides information on the top ten genes of the linear modeling, including their regulation status. Information on the top 200 such cancer-specific genes from the linear modelling are provided in Supplementary File S2. Figure 4 shows violin-plot representations of expression distribution of the top ranked genes of the linear model. Violin plots for all the top 200 genes from the linear model are provided in Supplementary File S3.

**Figure 3:**
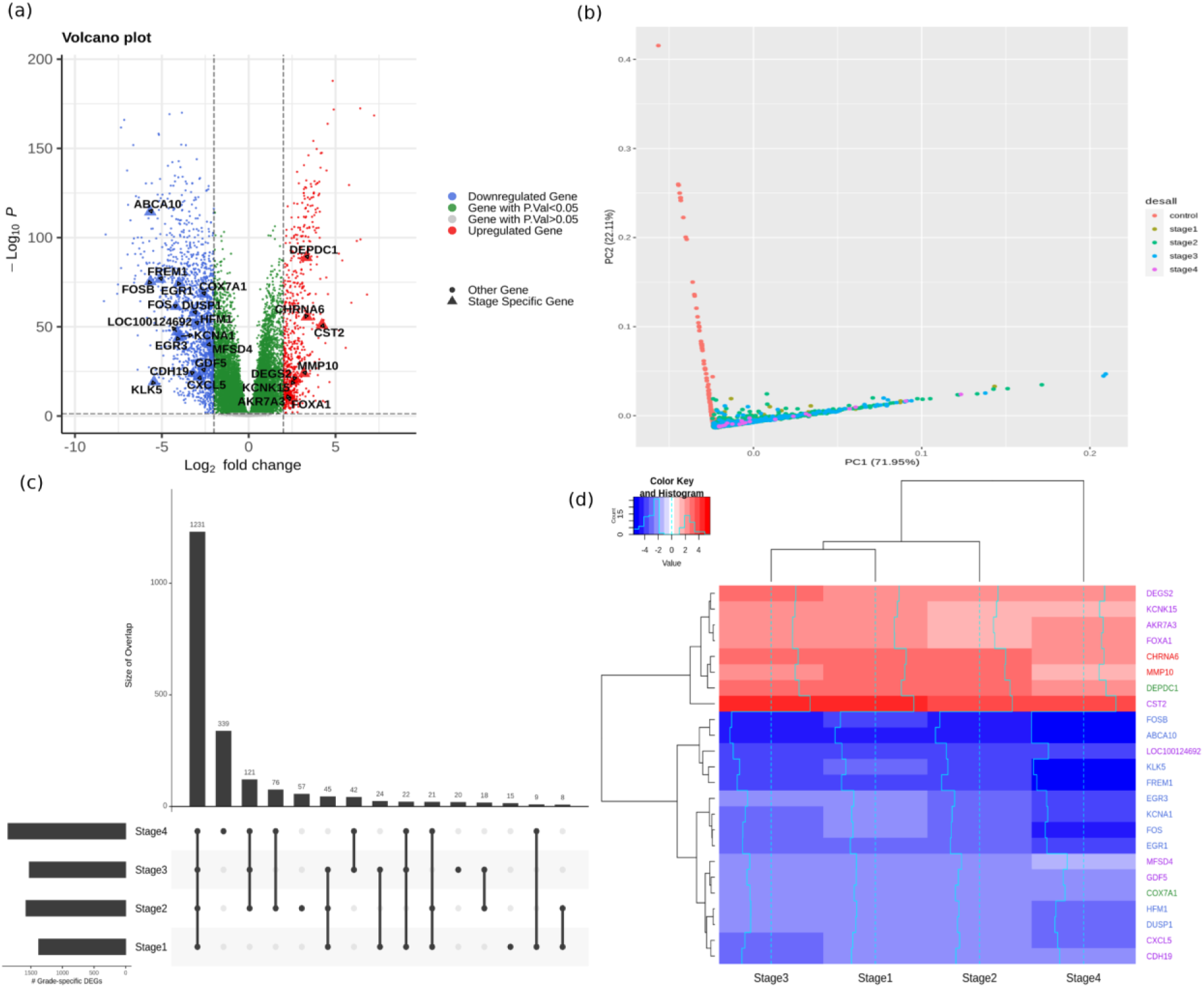
(a) Volcano plot of statistical significance vs log-fold change of differentially expressed genes. Downregulated genes (log-fold change < 2) are shown as blue dots, whereas upregulated genes (log-fold change > 2) are shown as red dots. Stage-salient genes are highlighted. (b) Top two principal components of the expression matrix of the top-ten genes from linear modelling. Normal samples can be seen to orient away from cancer samples. (c) A representation of differentially expressed genes to convey the trends across stages. (d) Heatmap representation of the stagewise expression of the 24 stage-salient genes, with both sample and gene dendrograms. It is seen that the gene dendrogram exhibits two main clusters, corresponding to overexpressed genes (red) and downregulated genes (blue). Euclidean distance was used as the metric for hierarchical clustering.

**Figure 4:**
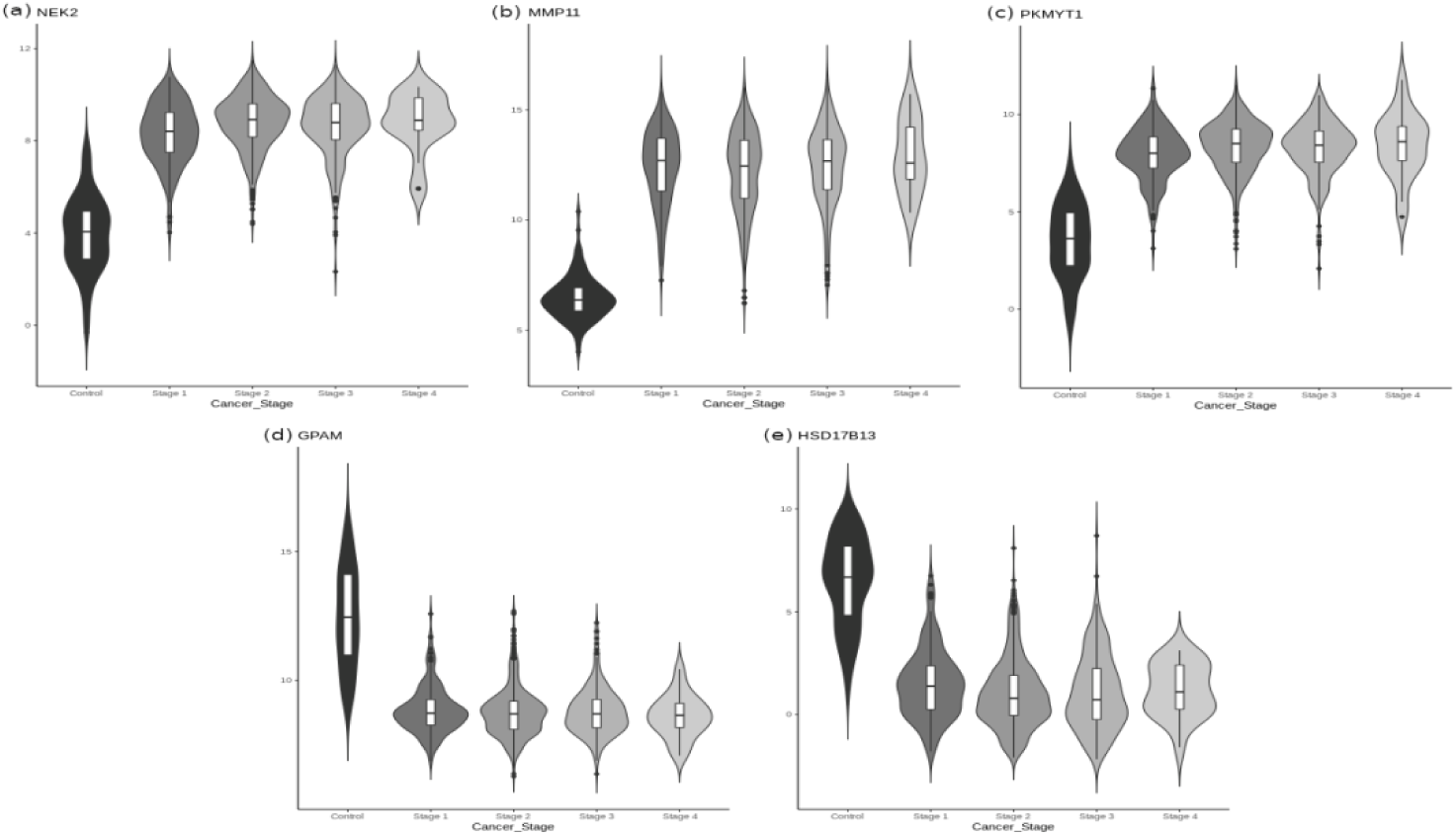
Distribution of expression of the top-ranked five genes in linear model, sorted by sample stage. It is seen that NEK2, MMP11 and PKMY11 (top row) are overexpressed in cancers, whereas GPAM and HSD17B13 are downregulated in cancers.

**Table 4.**
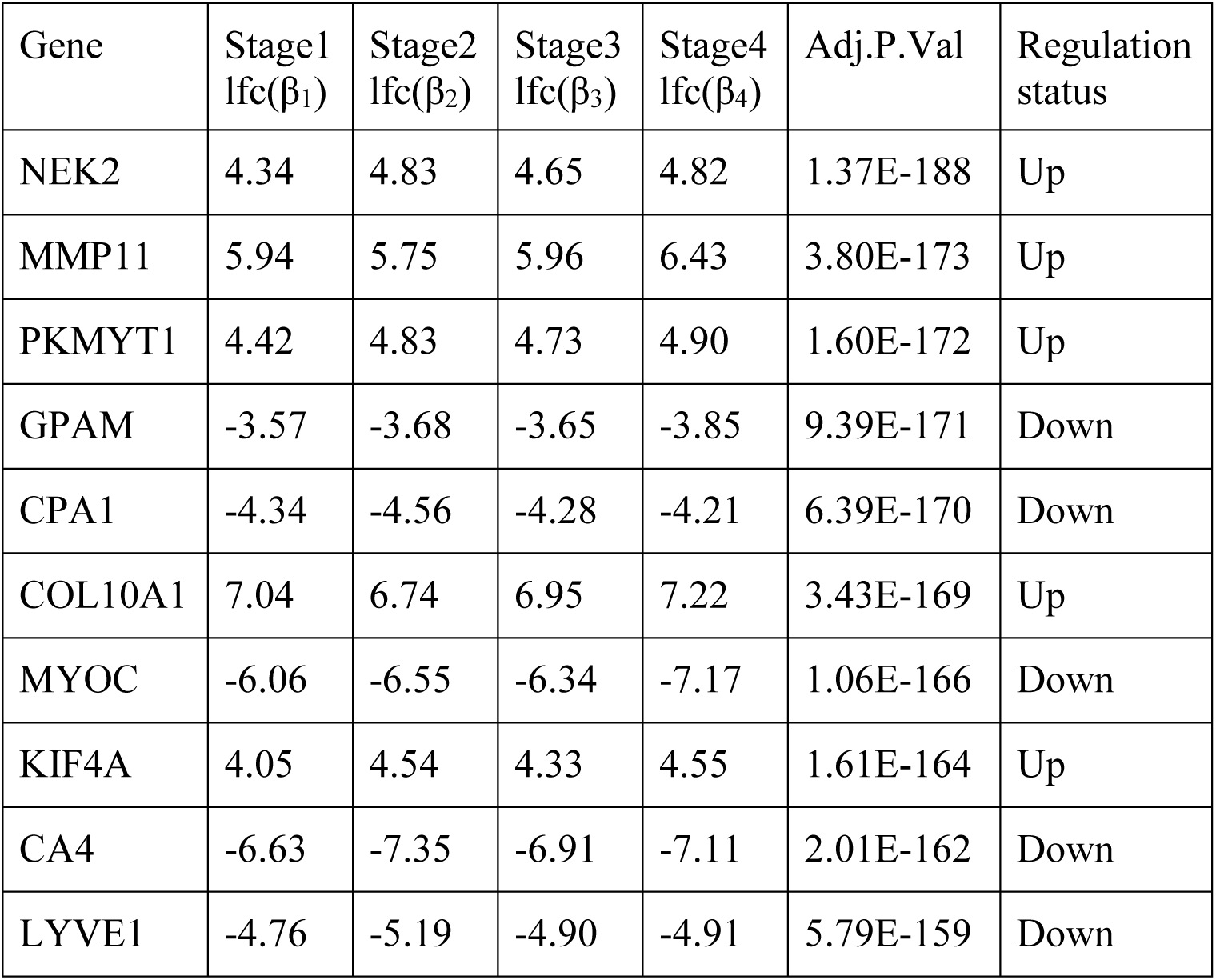
Top ten genes of the linear model with their stagewise mean log-fold change with respect to control. FDR-corrected significance and inferred regulation type are indicated.

| Gene | Stage1<br>lfc($\beta_1$) | Stage2<br>lfc($\beta_2$) | Stage3<br>lfc($\beta_3$) | Stage4<br>lfc($\beta_4$) | Adj.P.Val | Regulation<br>status |
| --- | --- | --- | --- | --- | --- | --- |
| NEK2 | 4.34 | 4.83 | 4.65 | 4.82 | 1.37E-188 | Up |
| MMP11 | 5.94 | 5.75 | 5.96 | 6.43 | 3.80E-173 | Up |
| PKMYT1 | 4.42 | 4.83 | 4.73 | 4.90 | 1.60E-172 | Up |
| GPAM | -3.57 | -3.68 | -3.65 | -3.85 | 9.39E-171 | Down |
| CPA1 | -4.34 | -4.56 | -4.28 | -4.21 | 6.39E-170 | Down |
| COL10A1 | 7.04 | 6.74 | 6.95 | 7.22 | 3.43E-169 | Up |
| MYOC | -6.06 | -6.55 | -6.34 | -7.17 | 1.06E-166 | Down |
| KIF4A | 4.05 | 4.54 | 4.33 | 4.55 | 1.61E-164 | Up |
| CA4 | -6.63 | -7.35 | -6.91 | -7.11 | 2.01E-162 | Down |
| LYVE1 | -4.76 | -5.19 | -4.90 | -4.91 | 5.79E-159 | Down |

Applying the level-I biomarker filters (|lfc| > 2 and p-value <0.001) yielded a total of 927 stage-specific genes (74 Stage-I, 238 Stage-II, 90 Stage-III, and 525 Stage-IV specific DEGs, visualized as an Upset plot^56^ in Figure 3c). For the identification of stage-salient genes two contrasts were applied with stringent criteria and the DEGs identified with different comparisons. This contrast has yielded 2 Stage I salient, 2 Stage II salient, 10 Stage III salient and 20 Stage IV salient genes. Limiting to the top ten stage-IV salient genes (by significance), we finally obtained 24 stage salient genes (Table 5). A heatmap visualization of the stage-salient genes exhibited a systematic differential regulation relative to the controls (Figure 3d). Stage III 4 genes cluster along with Stage I genes and DEPDC1 Stage II with outward CST2. Rest genes from stage III and stage IV form a cluster along with COX7A1 Stage II gene. Violin plots of expression distribution across sample phenotypes for these genes could be found in Supplementary File S4.

**Table 5:** Trends in mean expression of stage-salient genes with cancer progression. The inferred regulation status in cancer is noted.

| Gene | Stage Information | $\beta_0$ | $\beta_1$ | $\beta_2$ | $\beta_3$ | $\beta_4$ | Adj.P.Val<br>1<br>(from contrast) | Adj.P.Val<br>(from control) | Regulation status |
| --- | --- | --- | --- | --- | --- | --- | --- | --- | --- |
| CHRNA6 | Stage I | -1.67 | **3.35** | 2.85 | 2.93 | 2.21 | 2.25E-52 | 7.59E-51 | Up |
| MMP10 | Stage I | 0.04 | **3.19** | 2.76 | 2.61 | 1.68 | 5.07E-23 | 1.66E-24 | Up |
| DEPDC1 | Stage II | 2.01 | 2.83 | **3.32** | 3.03 | 2.43 | 3.26E-92 | 1.39E-89 | Up |
| COX7A1 | Stage II | 2.36 | -2.31 | **-2.62** | -2.30 | -2.03 | 3.15E-72 | 4.39E-69 | Down |
| KCNK15 | Stage III | 1.99 | 2.40 | 1.85 | **2.59** | 1.72 | 8.24E-21 | 5.27E-20 | Up |
| MFSD4 | Stage III | 1.56 | -2.06 | -1.96 | **-2.32** | -1.79 | 4.51E-41 | 2.88E-41 | Down |
| CDH19 | Stage III | -3.13 | -2.60 | -2.58 | **-3.19** | -2.61 | 3.31E-26 | 1.53E-24 | Down |
| CXCL5 | Stage III | -2.03 | -2.47 | -2.17 | **-2.87** | -2.83 | 5.12E-24 | 1.30E-22 | Down |
| AKR7A3 | Stage III | 3.26 | 2.05 | 1.52 | **2.33** | 2.12 | 1.83E-13 | 2.55E-12 | Up |
| DEGS2 | Stage III | 4.82 | 2.60 | 2.02 | **2.69** | 2.27 | 9.30E-22 | 1.68E-21 | Up |
| CST2 | Stage III | -0.60 | 4.18 | 3.57 | **4.22** | 3.52 | 2.19E-48 | 8.75E-52 | Up |
| LOC100124692 | Stage III | -2.52 | -3.64 | -3.60 | **-4.13** | -3.83 | 2.98E-46 | 8.24E-48 | Down |
| GDF5 | Stage III | -1.26 | -2.08 | -2.31 | **-2.63** | -2.24 | 1.67E-26 | 3.64E-26 | Down |
| FOXA1 | Stage III | 7.19 | 2.09 | 1.64 | **2.32** | 1.94 | 4.81E-13 | 1.30E-11 | Up |
| EGR3 | Stage IV | 4.14 | -2.33 | -2.71 | -2.57 | **-4.04** | 3.53E-18 | 1.46E-44 | Down |
| FOS | Stage IV | 7.27 | -2.44 | -3.07 | -3.09 | **-4.19** | 3.40E-21 | 3.50E-62 | Down |
| FOSB | Stage IV | 4.71 | -3.80 | -4.33 | -4.30 | **-5.66** | 9.16E-25 | 4.51E-76 | Down |
| DUSP1 | Stage IV | 7.00 | -2.13 | -2.40 | -2.23 | **-3.13** | 2.51E-19 | 1.81E-58 | Down |
| FREM1 | Stage IV | 0.85 | -3.67 | -4.13 | -3.70 | **-5.09** | 1.29E-23 | 2.43E-77 | Down |
| EGR1 | Stage IV | 7.45 | -2.72 | -3.18 | -3.11 | **-4.00** | 3.63E-23 | 2.23E-75 | Down |
| HFM1 | Stage IV | -3.44 | -2.02 | -2.24 | -2.23 | **-3.02** | 6.13E-18 | 1.43E-52 | Down |
| ABCA10 | Stage IV | -0.28 | -4.38 | -4.80 | -4.48 | **-5.67** | 5.63E-33 | 3.89E-115 | Down |
| KLK5 | Stage IV | 1.26 | -3.21 | -3.44 | -3.44 | **-5.45** | 6.93E-20 | 2.41E-09 | Down |
| KCNA1 | Stage IV | -1.69 | -2.58 | -2.99 | -2.81 | **-3.93** | 3.08E-15 | 1.99E-45 | Down |

The GO and KEGG pathway analysis was performed for the Stage salient genes to identify over-represented biological processes among these candidate biomarkers (complete results in Supplementary File S5 and S6, respectively). Genes that were monotonically expressed with cancer progression were identified by observing the trend in mean expression with increasing cancer stage. This yielded 2246 significantly monotonic genes (1015 with increasing expression, and 1231 with decreasing expression). The top 20 such genes with their inferred regulation status are shown in Table 6. A stage-specific gene is said to be contra-regulated when its mean expression is “paradoxical” with cancer progression. There are six patterns of “paradoxical” mean expression, enumerated in Supplementary File S7. We identified 112 stage-specific genes with such contra-regulation, including one stage-I salient gene (CHRNA6). Contra-regulated genes exhibit unstable expression with cancer progression, and their anomalous behavior might represent possible directions for experimental investigations (Supplementary File S7). Stage-specific DEGs devoid of such contra-regulation suggest a more general role as enhancers of cancer progression.

**Table 6:** Top 20 genes with significant monotonic patterns of expression. Intercept, coefficient and adj. p-values from the ordinal model are used. Status indicates monotonic upregulation (UP) or monotonic downregulation (DOWN). The table is sorted by significance (adj.p-value). Adj. R^2^ goodness-of-fit of a stage-ordinal model of expression for each gene is provided.

| Gene | Intercept | Coefficient | Adj.P-value | Adj.$R^2$ | Status |
| --- | --- | --- | --- | --- | --- |
| FAM13A | 9.842826 | -0.62121 | 1.70E-64 | 0.2255 | DOWN |
| GABRD | 3.697762 | 0.889287 | 2.27E-64 | 0.2249 | UP |
| KLHL31 | 6.778289 | -0.8667 | 2.33E-63 | 0.2217 | DOWN |
| POC1A | 6.587719 | 0.525973 | 4.14E-63 | 0.2209 | UP |
| PAFAH1B3 | 8.753896 | 0.602506 | 1.23E-62 | 0.2193 | UP |
| SORBS1 | 11.50753 | -0.83632 | 5.17E-62 | 0.2174 | DOWN |
| NIPSNAP3B | 6.082268 | -0.70387 | 1.27E-61 | 0.2161 | DOWN |
| TMEM220 | 6.96875 | -0.67023 | 7.56E-60 | 0.2102 | DOWN |
| SPTBN1 | 13.42746 | -0.45273 | 2.81E-59 | 0.2083 | DOWN |
| SIK2 | 10.23114 | -0.52331 | 2.56E-58 | 0.2051 | DOWN |
| RECQL4 | 6.916714 | 0.743136 | 1.59E-57 | 0.2025 | UP |
| C7orf41 | 10.91012 | -0.61324 | 1.81E-57 | 0.2023 | DOWN |
| RAG1AP1 | 9.736787 | 0.453142 | 5.56E-57 | 0.2001 | UP |
| HSD17B6 | 4.70826 | 0.715399 | 6.98E-57 | 0.2004 | UP |
| SLC35A2 | 9.380796 | 0.311207 | 7.48E-57 | 0.2002 | UP |
| CCDC64 | 6.871398 | 0.724435 | 3.72E-56 | 0.1979 | UP |
| DMD | 9.497599 | -0.92277 | 2.47E-55 | 0.1952 | DOWN |
| RUSC1 | 9.565741 | 0.353172 | 1.24E-53 | 0.1897 | UP |
| CXCL2 | 6.668874 | -1.23033 | 4.45E-53 | 0.1877 | DOWN |
| PRR19 | 4.794229 | 0.497467 | 1.87E-52 | 0.1857 | UP |

**Table 7:**
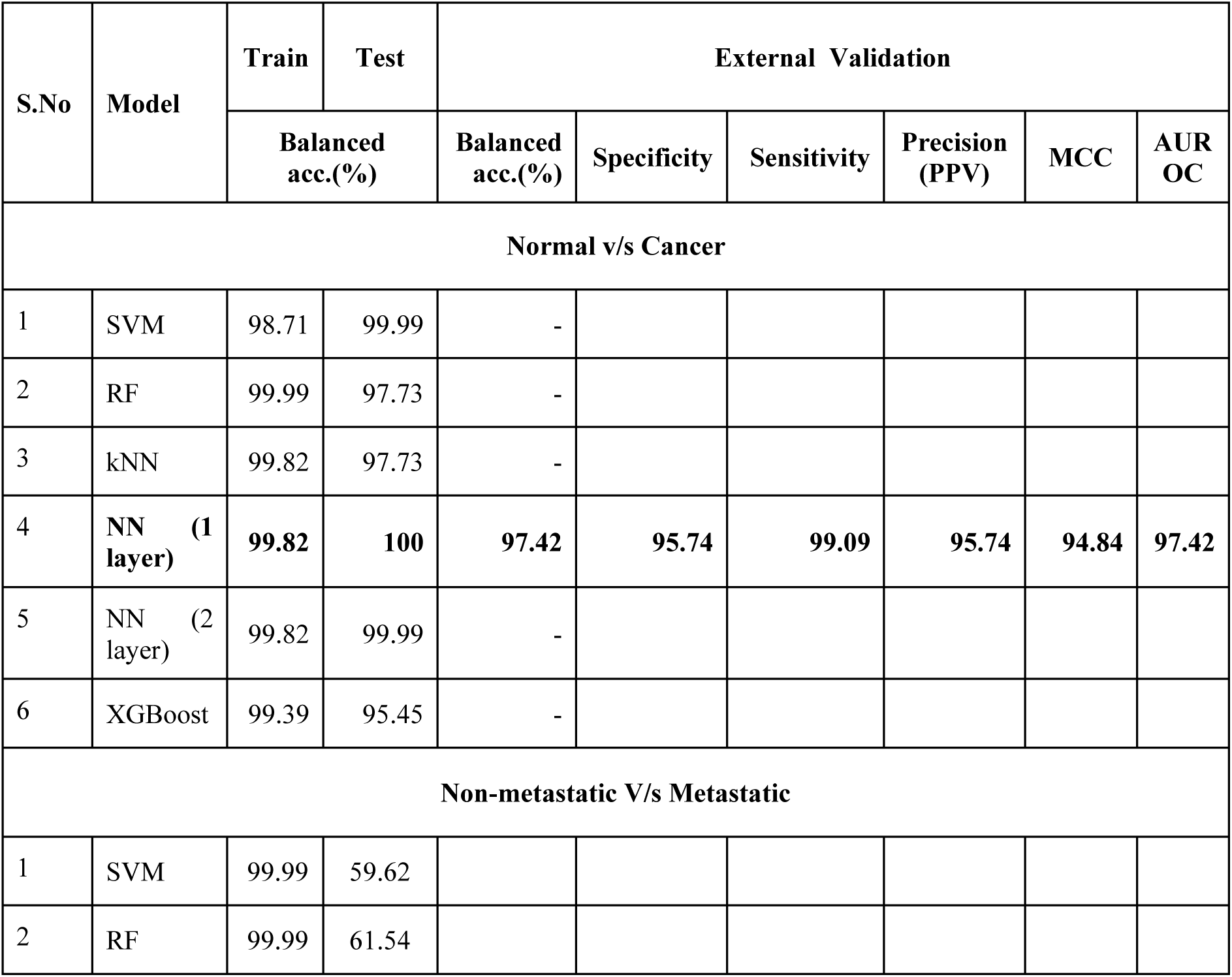
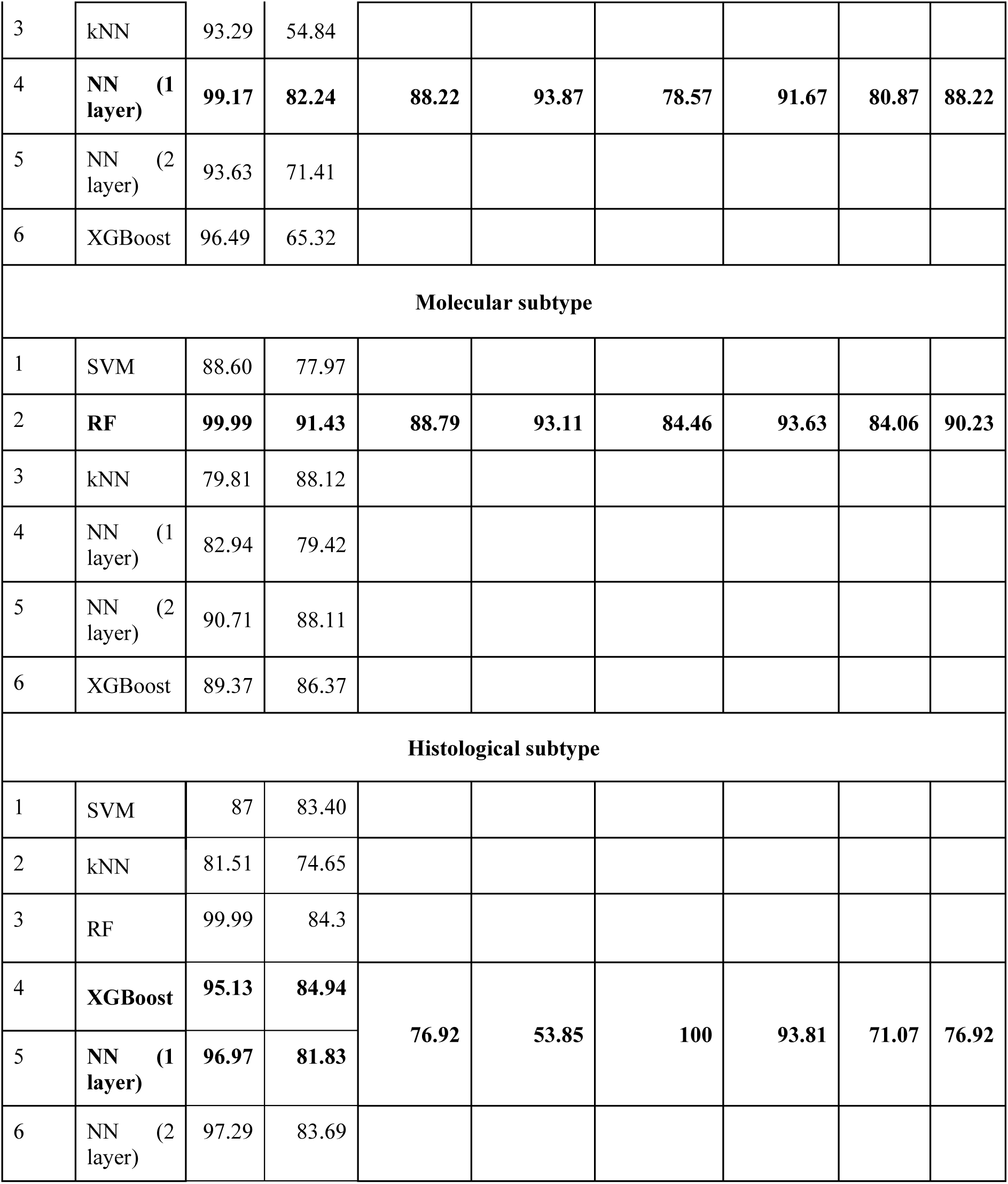
Performance of different ML models for the different problems. Four problems are discussed: (i) normal v/s cancer using ten features, (ii) metastatic v/s non-metastatic using five features, (iii) molecular subtyping using 16 features, and (iv) histological subtyping using 24 features. The model with the best performance (balanced accuracy) on the holdout testset for each problem is identified and externally validated. MCC and AUROC values of the best model are rescaled to the range [0,100].

### Normal v/s cancer

Stratified sampling of the TCGA BRCA dataset based on the class ‘cancer’ or ‘normal’ yielded a training dataset of 90 Normal and 854 Cancer samples, and a test dataset of 22 Normal and 212 Cancer samples. Completion of feature engineering yielded ten consensus features for model development, including two stage-salient genes (FREM1, ABCA10) and eight genes from the linear model (NEK2, MMP11, PKMYT1, GPAM, CPA1, COL10A1, CA4, LYVE1). Of the six different ML models trained, the neural network with one hidden layer model provided the best performance, yielding 99.82 % balanced accuracy on the training dataset and 100 % on the holdout testset (Table 8). The model was re-built using the full dataset and validated on external datasets: (i) with BRCA-KR, it yielded a balanced accuracy ∼ 94.00%; and (ii) with GTEx, the model produced an accuracy ∼ 100% (all correct predictions), yielded an overall balanced accuracy ∼ 97.42% on external validation. The details could be found in Supplementary File S8. Supplementary File S8 provides the prediction probabilities for all instances in the external validation as well. Prediction probability is a measure of the strength of evidence for the predicted class, and based on the distribution of its values, recommendations for evidence of the predicted class may be generated. Correct predictions were supported by very strong prediction probabilities (> 0.9) relative to incorrect predictions.

**Table 8:** Performance of histological subtype model on external validation. The confusion matrix of the ensemble model on brca_mbcproject_wagle_2017 is provided below. Instances with ambiguous predictions were omitted in evaluating the ensemble model.

| Confusion matrix |  | Ground Truth |  |
| --- | --- | --- | --- |
|  |  | Ductal | Lobular |
| Predicted | Ductal | 91 | 6 |
|  | Lobular | 0 | 7 |

### Non-metastatic v/s Metastatic

Stratified sampling of the TCGA BRCA dataset based on the class ‘non-metastatic’ or ‘metastatic’ yielded a training dataset of 837 non-metastatic and 16 Metastatic samples, and a test dataset of 209 non-metastatic and 4 Metastatic samples. SMOTE balancing of the training dataset yielded a dataset with 480 non-metastatic and 176 Metastatic samples. Completion of feature engineering yielded five consensus features for model development, namely DEPDC1, FOSB, DUSP1, MMP27 and ABCA10. Of the six different ML models trained, the neural network with one hidden layer model provided the best performance, yielding 99.14% balanced accuracy on the training dataset and 82.24% on the holdout testset (Table 8). The model was re-built using the full dataset and validated on the BRCA-KR and GSE18549 datasets, yielding an overall balanced accuracy ∼ 88.22% on the external validation. The details could be found in Supplementary File S9. Supplementary File S9 provides the prediction probabilities for all instances in the external validation. On inspection of the distribution of prediction probabilities, correct predictions were found to be supported by high values (> 0.75) relative to incorrect predictions.

### Molecular Subtype classification

Stratified sampling of the TCGA BRCA dataset based on the molecular subtype class (‘Luminal’ or ‘TNBC’ or ‘HER2’) yielded a training dataset of 434 Luminal, 30 HER2 and 92 TNBC samples, and a test dataset of 113 Luminal, 7 HER2 and 23 TNBC samples. Completion of feature engineering yielded 16 consensus features for model development, namely GATA3, AGR3, CA12, TBC1D9, ERBB2, MLPH, KCNK15, ANXA9, FLJ45983, GRB7, PGAP3, STARD3, SLC44A4, PCSK6, FOXA1 and BCAS1. Of the six different ML models trained, the Random forest model provided the best performance, yielding 100% balanced accuracy on the training dataset and 91.43% on the holdout testset (Table 8). The model was re-built using the full dataset and was validated with external datasets. Validation with the METABRIC dataset yielded a balanced accuracy ∼ 88.79%. Validation on out-of-chort TNBC-only dataset yielded a single misclassification (Luminal) out of 26 TNBC samples. The details could be found in the Supplementary File 10. Supplementary File S10 and S12 provide the prediction probabilities for all instances in the external validation (METABRIC and TNBC datasets, respectively). On inspection of the distribution of prediction probabilities, correct predictions were found to be supported by high values (> 0.7) relative to incorrect predictions. In addition, we investigated the RandomForest model for feature importance using R caret^57^. Feature importance was measured based on mean decrease in Gini score, and the top five features were GATA3, CA12, AGR3, TBC1D9, and MLPH (Figure 5).

**Figure 5:**
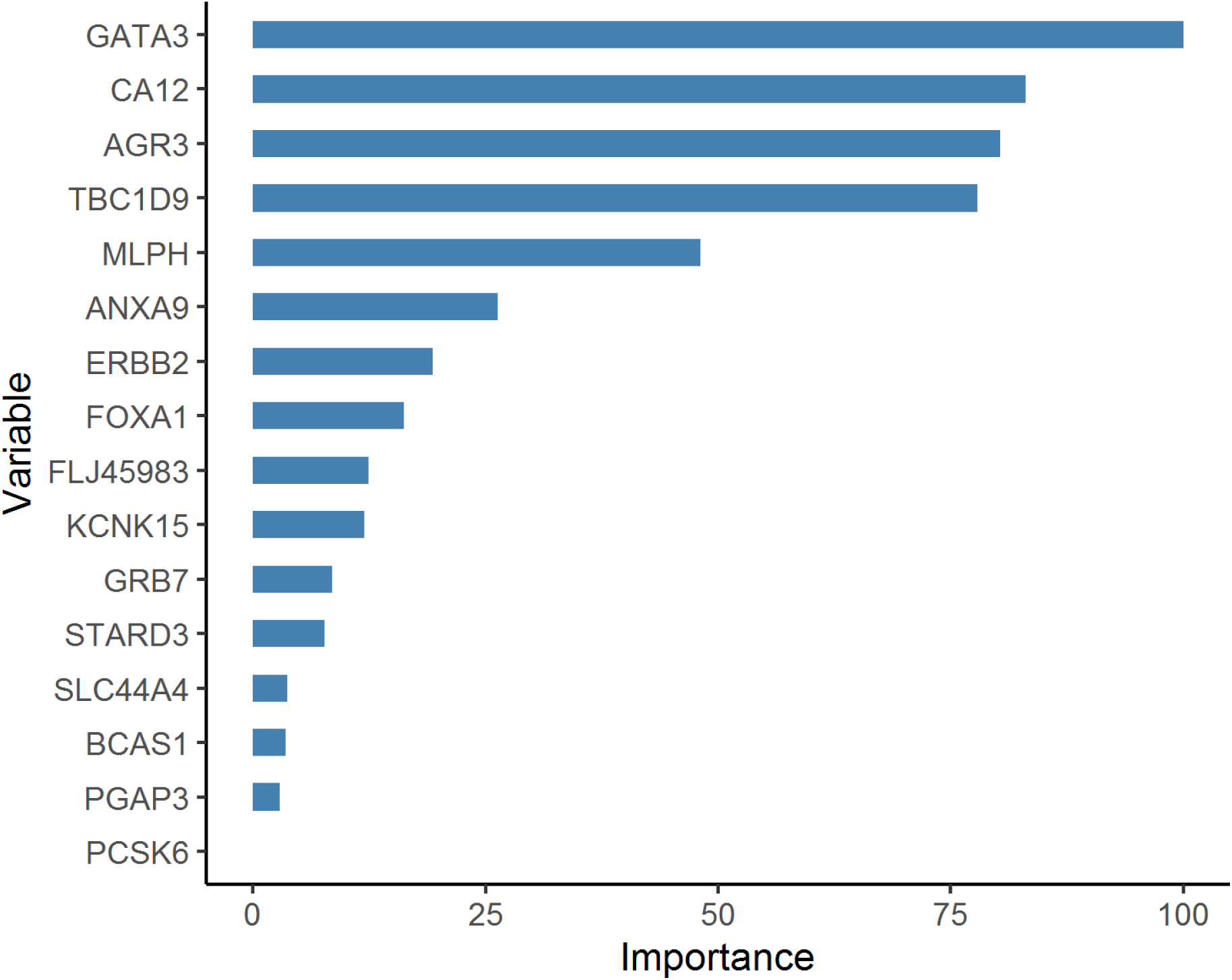
Importance ranking of features used in developing the molecular subtype model. The scores are normalized with respect to the top-scoring feature, GATA3.

### Histological Subtype classification

Stratified sampling of the TCGA BRCA dataset based on the histological subtype (‘IDC’ or ‘ILC’) yielded a training dataset of 624 IDC and 162 ILC samples, and a test dataset of 156 IDC and 40 ILC samples. A contrast of ductal and lobular samples followed by the completion of feature engineering yielded 24 final features for model development, namely ADCY5, ALDH1L1, ANKRD43, C1orf64, C7, CAPN8, CCL14, CDH1, CIDEA, CTSG, DARC, F7, FXYD1, HPX, IGFN1, MMP1, PEBP4, PLCXD3, PROL1, SHROOM1, TFAP2B, TFF1, TNNT3, and WNK4. Of the six different ML models trained, the models with >95% balanced accuracy on the training dataset were identified and bundled into an ensemble classifier. This included the XGBoost model and the neural network with 1-layer model, both yielding 88.74% balanced accuracy on the holdout testset (Table 8). The ensemble model re-built using the full dataset was validated on the external dataset: brca_mbcproject_wagle_2017, encoding both the histological subtypes of interest (IDC and ILC) as well as other subtypes such as ‘mixed histology’, ‘DCIS’ (ductal carcinoma in situ), and ‘NOS’. If the constituent models of the ensemble disagreed on the predicted class, then the ensemble classifier is challenged. Inconsistent ensemble predictions might be instructive, but need further disambiguation. Omitting the eleven such instances from the external dataset, we obtained ensemble accuracy ∼94.23% and balanced accuracy ∼76.92% (Table 8). It is clear that the ensemble model has encountered generalization errors in learning the ILC class. The details could be found in Supplementary File 11. Supplementary File S11 provides the prediction probabilities for all instances in the external validation. On inspection of the distribution of prediction probabilities, correct predictions were found to be supported by high values (> 0.7) relative to incorrect predictions.

### Validation with miRNA analysis

Stage-salient miRNA were identified using the two-level contrasts of the miRNA expression data, and then their targets were identified using the R multiMiR package (Supplementary File S12). Based on these results, we determined the concordance between the regulatory miRNAs and their target genes. Temporal concordance in expression exists if the salience in miRNA expression is at least as early as the salience in target gene expression. If the expression pattern of an miRNA is antagonistic to that of its target gene, a paradoxical aberration with a protective function is possible. Table 9 summarizes the validation of stage-salient gene expression from the angle of miRNA expression. Concordance between the mRNA and miRNA in both direction of expression and temporal dimension is achieved for 13 stage-salient genes: MMP10, DEPDC1, CDH19, FOXA1, DEGS2, CST2, AKR7A3, EGR1, EGR3, FOS, FOSB, FGF2, and HCN2. Among the stage-salient genes that feature in the ML models, DEPDC1 and FOSB are strongly-supported as candidate biomarkers. We also sought to identify the key regulatory miRNAs decoded by stage. In summary, 25 stage-salient miRNAs were identified (Supplementary File S12), and found to regulate most of the stage-salient genes. Stage-salient miRNA that were fully concordant with target mRNAs included hsa-miR-182-5p, hsa-miR-210-3p, hsa-miR10b-5p, hsa-miR-200a-5p, hsa-miR-96-5p, hsa-miR-21-5p, hsa-miR-133a-3p, hsa-miR-335-5p, hsa-miR-204-5p, and hsa-miR-145-5p. Further, four of the stage-salient miRNAs regulated genes that featured in the ML models, namely hsa-miR-210-3p, hsa-miR10b-5p, hsa-miR-200a-5p, and hsa-miR-96-5p. Only five stage-salient miRNAs displayed targets none of which were a stage-salient gene, and conversely, eleven stage-salient genes were not subject to regulation by a stage-salient miRNA (namely COX7A1, DACT2, KCNK15, MFSD4, DSC3, KLK5, KRT15, LOC100124692, ABCA10, MAPK8IP2, and MASP1). The detailed analysis is provided in Supplementary File S12.

**Table 9:** Putative target stage-salient genes mapped with their regulatory stage-salient miRNA. Concordance in expression is noted if miRNA overexpression is observed with target gene downregulation or vice-versa. Evaluation of temporal concordance is useful if concordance in expression exists. If there is no concordance in expression, temporal concordance is not evaluated. Genes that display concordance with regulatory miRNA in the direction of expression as well as temporal dimension are **emphasized**. Target stage-salient genes that represent features used in the ML models are *italicized*. Upregulated miRNAs denote candidate oncomiRs, whereas downregulated miRNAs denote candidate TSmiRs.

| S.No | Gene |  |  | Regulatory miRNA |  |  |
| --- | --- | --- | --- | --- | --- | --- |
|  | Name | Expression | Salience | Name | Concordance |  |
|  |  |  |  |  | Expression | Temporal |
| 1 | CHRNA6 | Up | Stage I | hsa-miR-452-3p | Yes | No |
| 2 | **MMP10** | **Up** | **Stage I** | **hsa-miR-182-5p** | **Yes** | **Yes** |
|  |  |  |  | hsa-miR-210-3p | Yes | No |
| 3 | **DEPDC1** | **Up** | **Stage II** | **hsa-miR-200b-3p** | Yes | Yes |
|  |  |  |  | **hsa-miR-210-3p** | Yes | Yes |
|  |  |  |  | **hsa-miR10b-5p** | Yes | Yes |
|  |  |  |  | **hsa-miR-200a-5p** | Yes | Yes |
|  |  |  |  | **hsa-miR-96-5p** | No | - |
| 4 | **CDH19** | **Down** | **Stage III** | **hsa-miR10b-5p** | No | - |
|  |  |  |  | **hsa-miR-182-5p** | No | - |
|  |  |  |  | **hsa-miR-335-5p** | No | - |
| 5 | **GDF5** | **Down** | **Stage III** | **hsa-miR-21-5p** | Yes | No |
|  |  |  |  | **hsa-miR-335-5p** | No | - |
|  |  |  |  | **hsa-miR-182-5p** | No | - |
| 6 | **FOXA1** | **Up** | **Stage III** | **hsa-miR-200a-3p** | Yes | Yes |
|  |  |  |  | **hsa-miR-141-3p** | No | - |
| 7 | **DEGS2** | **Up** | **Stage III** | **hsa-miR-200b-3p** | Yes | Yes |
| 8 | **CST2** | **Up** | **Stage III** | **hsa-miR-210-3p** | Yes | Yes |
|  |  |  |  | **hsa-miR-335-5p** | Yes | No |
| 9 | **AKR7A3** | **Up** | **Stage III** | **hsa-miR-210-3p** | Yes | Yes |
| 10 | **CXCL5** | **Down** | **Stage III** | **hsa-miR10b-5p** | No | - |
| 11 | **EGR1** | **Down** | **Stage IV** | **hsa-miR-21-5p** | Yes | Yes |
|  |  |  |  | **hsa-miR183-5p** | Yes | Yes |
|  |  |  |  | **hsa-miR-204-5p** | No | - |
|  |  |  |  | hsa-miR-133a-3p | No | - |
|  |  |  |  | hsa-miR-452-5p | No | - |
|  |  |  |  | hsa-miR-224-5p | No | - |
|  |  |  |  | hsa-miR10b-5p | No | - |
|  |  |  |  | hsa-miR-210-3p | No | - |
|  |  |  |  | hsa-miR-182-5p | No | - |
| 12 | **EGR3** | **Down** | **Stage IV** | **hsa-miR183-5p** | Yes | Yes |
|  |  |  |  | hsa-miR-335-5p | No | - |
|  |  |  |  | hsa-miR10b-5p | No | - |
|  |  |  |  | hsa-miR-182-5p | No | - |
| 13 | **FOSB** | **Down** | **Stage IV** | **hsa-miR183-5p** | Yes | Yes |
|  |  |  |  | hsa-miR-224-3p | No | - |
|  |  |  |  | hsa-miR-224-5p | No | - |
|  |  |  |  | hsa-miR-200b-3p | No | - |
| 14 | **KLK7** | **Down** | **Stage IV** | hsa-miR-335-5p | No | - |
|  |  |  |  | hsa-miR-182-5p | No | - |
| 15 | **DUSP1** | **Down** | **Stage IV** | hsa-miR10b-5p | No | - |
|  |  |  |  | hsa-miR-200b-3p | No | - |
|  |  |  |  | hsa-miR-200b-3p | No | - |
| 17 | **FOS** | **Down** | **Stage IV** | **hsa-miR-196a-5p** | Yes | Yes |
|  |  |  |  | **hsa-miR183-5p** | Yes | Yes |
|  |  |  |  | hsa-miR-335-5p | No | - |
|  |  |  |  | hsa-miR10b-5p | No | - |
|  |  |  |  | hsa-miR-139-5p | No | - |
|  |  |  |  | hsa-miR-182-5p | No | - |
| 18 | KCNA1 | Down | Stage IV | hsa-miR-210-3p | No | - |
| 19 | **FGF2** | **Down** | **Stage IV** | **hsa-miR-196a-5p** | Yes | Yes |
|  |  |  |  | **hsa-miR-96-5p** | Yes | Yes |
|  |  |  |  | hsa-miR-145-5p | No | - |
|  |  |  |  | hsa-miR-133a-3p | No | - |
|  |  |  |  | hsa-miR10b-5p | No | - |
|  |  |  |  | hsa-miR-210-3p | No | - |
|  |  |  |  | hsa-miR-182-5p | No | - |
| 20 | **HCN2** | **Up** | **Stage IV** | **hsa-miR-133a-3p** | Yes | Yes |
| 21 | KIT | Down | Stage IV | hsa-miR-335-5p | No | - |
| 22 | *FREM1* | *Down* | *Stage IV* | *hsa-miR-335-5p* | No | - |
| 23 | HFM1 | Down | Stage IV | hsa-miR-335-5p | No | - |

### Validation with methylation analysis

Aberrant methylation in the core / proximal promoter regions as well as enhancers could have profound regulatory effects on gene expression. We obtained a total of 22 stage-salient DMGs from the consensus of Averep, Mvalue, and MethylMix procedures: 1 stage-I salient DMG (VOPP1), 8 stage-II salient DMGs (HS3ST3B1, CPLX1, EGR1, GMDS, ITPKB, TGFB1I1, C6orf145, SHC1), 10 stage-III salient DMGs (BTLA, TNFAIP2, PHYHIPL, LYN, MAML2, C16orf62, GPRC5B, CAPN9, AIPL1, AGAP1), and 4 stage-IV salient DMGs (CNP, TSPYL5, SLC7A5, HCN2). Salient methylation of a gene is an epigenetic mechanism to tune gene expression, and will precede changes in its expression. In this respect, the stage-II salient methylation of EGR1 possibly set the stage for its stage-IV salience in expression. It is observed that the stage-IV salient hypermethylation of HCN2 was at odds with its stage-IV salient overexpression.

Mining the methylation patterns of all stage-salient genes for differential methylation-driven genes revealed five transcriptionally predictive genes negatively correlated with gene expression, namely AKR7A3, COX7A1, DEGS2, EGR1, and FOXA1 (Figure 6). Four of these genes exhibited two-component mixtures of methylation distribution, indicating a probable shift in methylation levels in cancer samples relative to healthy ones. COX7A1 showed three-component mixtures of methylation distribution, with an additional methylation component signifying cancer progression. Table 10 summarizes the information for each of these genes. FOXA1 showed a maximum inverse correlation of –0.66. Methylation of FOXA1 is concordant with the expression of its regulatory miRNA, but FOXA1 mRNA expression is at odds with its epigenetic profiles, suggesting that epigenetic modulation was being used to restore FOXA1 aberrant expression. Methylation of AKR7A3, DEGS2, EGR1, and COX7A1 is concordant with their mRNA expression, providing strong support for their stage-salience. In addition, the above genes except COX7A1 were modulated by stage-salient miRNAs and in a concordant manner. This suggests a concert between the different layers of omics, adding ‘definiteness’ to gene expression on the path to phenotypic states. The mixture decomposition of methylation patterns of the remaining stage-salient genes is provided in Supplementary File S13. It could be seen, for e.g., that the methylation of ABCA10 is positively correlated with its expression, escaping clear interpretation.

**Figure 6.**
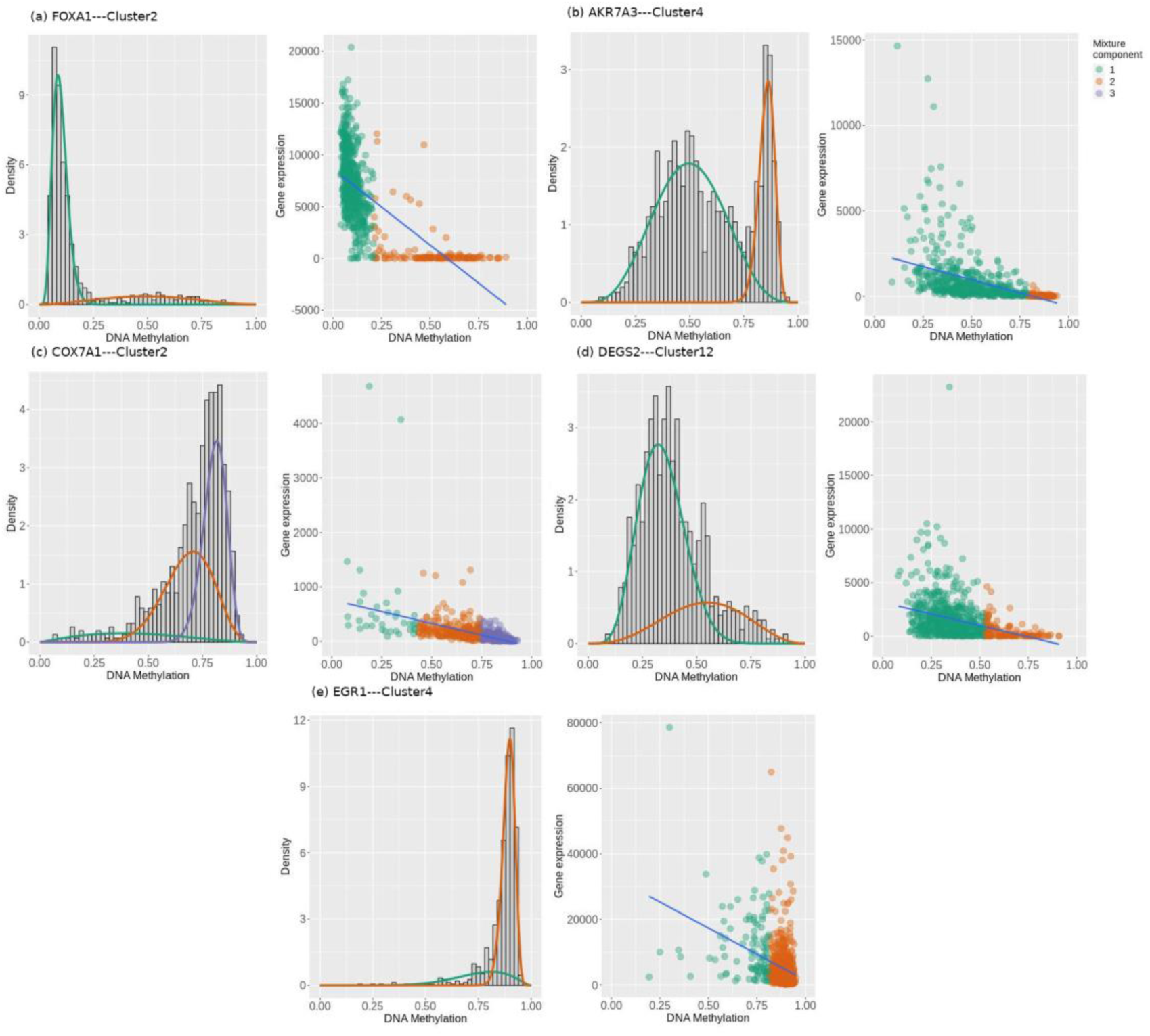
Mixture model of methylation pattern, and scatter of expression vs methylation for the respective cluster of each stage-salient differential methylation-driven gene. (a) AKR7A3 (b) COX7A1 (c) DEGS2 (d) EGR1 and (e) FOXA1. Two mixture components each are seen for AKR7A3, DEGS2, EGR1, and FOXA1, and three for COX7A1. Bayesian Information Criterion was used for estimating the number of mixture components. A strong inverse correlation between expression and methylation is apparent for FOXA1. Visualized using MethylMix

**Table 10:** Summary of the stage-salient differential methylation-driven genes. Since the methylation of each gene was assayed at a variable number of CpG probe locations, the methylation patterns at different probes for a given gene were clustered based on Pearson’s correlation coefficient cut-off (> 0.7). Significant clusters were used to obtain the rest of the values in the table (namely effect size of differential methylation across mixture components, significance of the methylation pattern, coefficient of correlation between expression and methylation, and concordance). Sign of the DM effect signifies the type of aberrant methylation (hyper / hypo) across the mixture components.

| Gene of interest | CpG sites |  | Significant cluster |  | DM effect size | p-value | Type of DM | Correlation with expression | Concordance |
| --- | --- | --- | --- | --- | --- | --- | --- | --- | --- |
|  | Probes | Clusters | ID | Probes |  |  |  |  |  |
| FOXA1 | 18 | 10 | Cluster2 | 5 | 0.373 | 1.25E-98 | Hyper | -0.66 | No |
| AKR7A3 | 14 | 6 | Cluster4 | 1 | -0.321 | 9.89E-48 | Hypo | -0.49 | Yes |
| COX7A1 | 4 | 2 | Cluster2 | 3 | 0.413 | 3.36E-45 | Hyper | -0.48 | Yes |
| DEGS2 | 15 | 13 | Cluster12 | 1 | -0.157 | 1.56E-25 | Hypo | -0.36 | Yes |
| EGR1 | 13 | 11 | Cluster4 | 2 | 0.185 | 1.21E-23 | Hyper | -0.35 | Yes |

## Discussion

Validation of the models implied that the identity of cancer biomarkers for these problems might be invariant of ethnicity, and suggested that our models are robust to out-of-domain test data, notwithstanding shifts in expression profiles. Recently, we applied dimensionality reduction and unsupervised learning to the space of nine expression features (viz. NEK2, PKMYT1, MMP11, CPA1, COL10A1, HSD17B13, CA4, MYOC, LYVE1) and addressed the ‘cancer’ vs. ‘normal’ binary classification, producing BrcaDx (https://apalania.shinyapps.io/BrcaDx)^58^ with a balanced accuracy of 95.52% on the BRCA-KR and GTEx. Here we have used a supervised learning approach to the same problem (Fig. 2), and derived ten features, including ABCA10, GPAM, FREM1, and the first seven features in the above-noted prior work. This has yielded a balanced accuracy of 97.42% on the same external datasets, which is a significant improvement. Beyond the performance improvement, it is noted that BrcaDx also suffers from the relative opaqueness of surrogate biomarker spaces that tend to obscure interpretation. Other recent advances for discriminating breast cancer from normal samples include a supervised learning model of 20 biomarkers, which was validated on only an internal test set with a balanced accuracy that does not exceed 86%^59^. BC-Predict and BrcaDx are both reproducible and interestingly share no common biomarkers with these earlier models.

### Literature discussion

We searched Pubmed (www.pubmed.gov) using the keyword: “breast cancer” AND “stage specific” AND “gene”, and found a handful of known stage-specific genes. TIEG (or KLF10) is an anti-metastasis / tumor-suppressor gene, which inhibits invasive breast cancer by blocking EGFR transcription in the EGFR signalling pathway^60^. Stage-specific expression of KLF10 in breast cancer biopsies has been published, with sustained downregulation leading to complete absence of expression in invasive subtypes^61^. Here KLF10 expression is found to be decreasing with stage relative to the normals. γ-Synuclein (SNCG) expression is strongly correlated with the stages of breast cancer, showing little expression in normal or benign samples and increasing expression with cancer stage, and detectable only in a subset of patients^62^. Here we find increasing expression of SNCG in late-stage cancers, but downregulated with respect to expression in normal samples, which is a contrarian finding.

### Top genes from linear models

Players in cell cycle regulation featured among the top genes of the linear model, namely NEK2, PKYMT1, DEPDC1, KIF4A and CA4. Aberrations in cell cycle regulation facilitate sustained proliferative signalling and evasion of the growth suppressor, which are complementary hallmarks of cancers^4^. The top 200 linear model genes were screened against the known cancer driver genes in Cancer Gene Census, yielding four hits: BUB1B, EBF1, PPARG, and RECQL4. RECQL4 is a key DNA helicase, with a vital role in the maintenance of genomic stability^63^. It has been found to be mutated and often upregulated in breast cancer^64^, and its tumor-promoting activity has been observed in sporadic breast cancers with aggressive tumor behavior^65^. Searching the top 200 MEGs against the Cancer Gene Census yielded two other hits: EGFR and QKI. EGFR is the first antitumor target to be identified, and known to be overexpressed in most of the TNBC and inflammatory breast cancers^66^, but associated with paradoxical function in metastatic cancer progression^67^. Significant downregulation of QKI has been noted in breast cancer relative to normal tissues, along with poor prognosis, which suggest its tumor-suppressor role^68^. Expression of SLUG and QKI was correlated with epithelial to mesenchymal transition (EMT), and showed promise for use in breast cancer prognosis^69^. Intersection of the top 200 linear model genes with the top 200 MEGs yielded 18 genes (including RECQL4), whereas intersection with the top 200 of the second linear model yielded 32 genes. We found 17 genes in common to all the three sets, including FAM13A, GABRD, and SORBS1. Supplementary File S14 presents the complete results. FAM13A is a hypoxia-induced gene in non-small lung cancer, increasing susceptibility to BC in a population-based cohort^70^. Genes coexpressed with GABRD in colon cancer showed an enrichment for breast cancer and HPV infection pathway^71^, hinting at a possible regulatory role for the monotonic expression of GABRD. Downregulation of SORBS1 in cancer samples was associated with increased metastasis and poor survival outcomes^72^. Stage-wise distribution of expression of some consensus genes is presented in Supplementary File S15.

The 34 stage-salient candidate biomarkers identified here were cross-referenced with the Human Protein Atlas^73^. We found 11 genes (2 stage-III salient genes and 9 stage-IV salient genes) annotated as ‘cancer related genes’, of which two stage-IV salient markers, namely EGR3 and KRT15, were specifically noted as prognostic markers of breast cancer (Supplementary File S16).

### Early-stage salient Genes (Figure 7)

Nicotine in tobacco exerts its action through nicotinic acetylcholine receptors, which initiate cell proliferation^74^, concording with the identification of CHRNA6 (neuronal nicotinic acetylcholine receptor) as stage-I salient here. The downregulation of CHRNA6 with cancer progression is supported by studies on nicotinic expression in non-small cell lung cancer progression, where expression of CHRNA6 was found higher in non-smokers than smokers^75^. MMP10 is a member of the peptidase M10 family of matrix metalloproteinases, and could set the stage for cancer progression by facilitating tumor cell dissociation, augmenting migration/invasion capability, promoting endothelial cell tube formation, and inducing the expression of key angiogenic and metastatic factors^76^. Recently, Piskor *et al.* proposed that MMP10 in combination with MMP3 and CA-15 could be used as a biomarker panel for early-stage BC through a non-invasive approach^77^. Both these results accord with maximum expression of MMP10 in the early stages of cancer, reaffirming the effectiveness of our study design in identifying stage-salient markers. DEPDC1 is a novel cell cycle gene regulating apoptosis^78^, whose over-expression signifies cancer progression in BC and its subtypes^79,80^. Here we have pinpointed the stage-II salience of DEPDC1 over-expression. COX7A1 is involved in mitochondrial metabolism and was identified as a tumor suppressor in invasive breast carcinoma, due to aberrant promoter hypermethylation^81^. The stage-II salience of COX7A1 obtained in our studies supports its further exploration as a new biomarker and therapeutic target.

**Figure 7:**
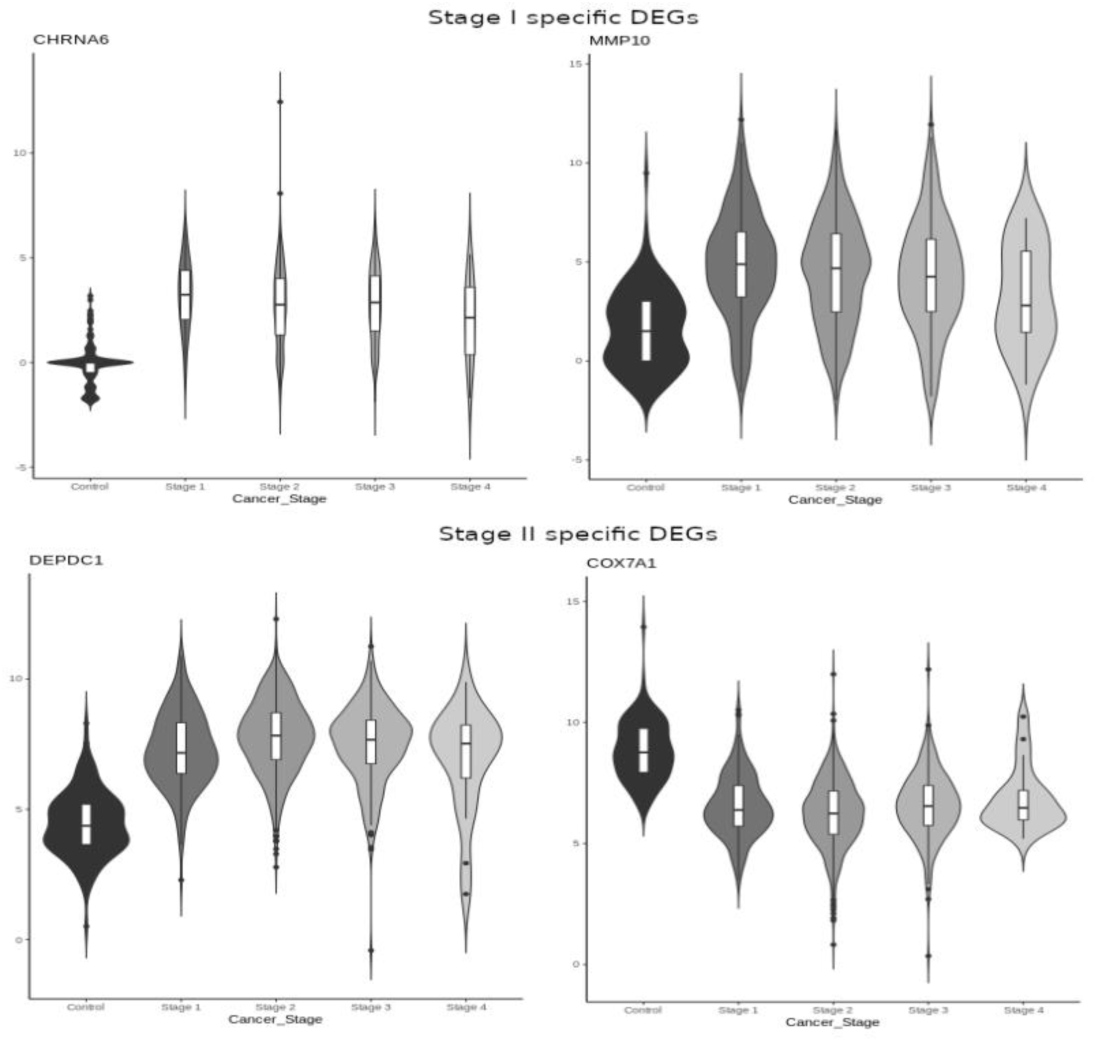
Distribution of expression patterns of stage-I and stage-II salient DEGs. CHRNA6 and MMP10 are both upregulated stage-I salient DE genes. With respect to the stage-II salient genes, DEPDC1 is upregulated and COX7A1 is downregulated.

### Stage III salient Genes (Figure 8)

It is known that KCNK15 is overexpressed in BC^82^, specifically in Luminal A subtype, but downregulated in TNBC subtype^83^. MFSD4 (major facilitator superfamily domain containing 4) has been identified as a tumor suppressor of cell motility and invasiveness (by influencing promoter methylation) and a biomarker of hepatic metastasis in gastric cancer^84^, correctly identified here as downregulated. CDH19 encodes a cell-cell adhesion receptor cadherin, essential to maintenance of intercellular connections, whose loss of function was observed in BC samples^85^. Aligning with this result, CDH19 is seen here to be downregulated. CXCL5, a chemokine, was found to regulate bone colonization in metastatic BC via its functional target CXCR2^86^, and its downregulation here might need further review. Oncogenic expression of AKR7A3 in the late stages of BC is detrimental to the period of disease-free survival, and it is interesting to note its stage-III salient upregulation here^87^. DEGS2 (delta(4)-desaturase sphingolipid 2) exhibits oncogenic expression in response to increased levels of ceramide in BC^88^, which resonates with the findings here. Growth differentiation factor-5 (GDF5) regulates TGFβ-mediated pro-angiogenic signaling^89^, and its significant downregulation in the late stages here might set the stage for metastatic cancer. Oncogenic expression of FOXA1 (Forkhead box A1) enables widespread epigenetic reprogramming in ER metastatic BC^90^, concordant with its overexpression here. Oncogenic expression of CST2 has been documented to promote bone metastasis in breast cancer^91^, borne out by its upregulated stage-III salience here.

**Figure 8:**
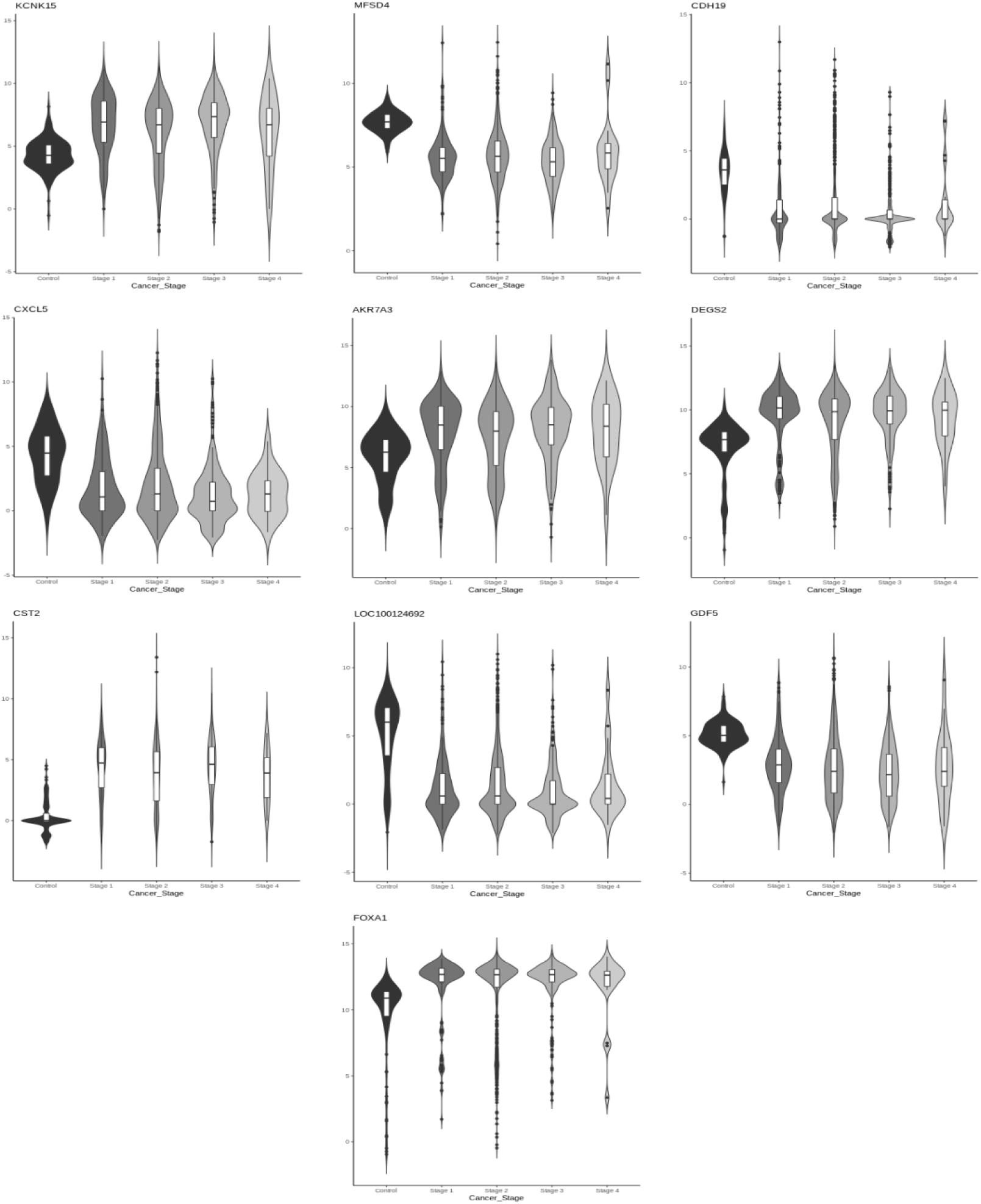
Distribution of expression patterns of stage-III salient genes. The mean expression of KCNK15, AKR7A3, DEGS2, CST2, and FOXA1 are smoothly upregulated, reaching a maximum in Stage-III. Conversely, the mean expression of MFSD4, CDH19, CXCL5, LOC100124692, and GDF5 DEGs are smoothly downregulated, reaching a minimum in stage-III.

### Stage IV salient Genes (Figure 9)

Suzuki *et al.* examined the role of EGR3 in BC and concluded that its overexpression in concert with the expression of other genes is necessary to establish invasive and metastatic BC^92^, which is in contradiction to the consistent downregulation seen here. FOS and FOSB showed near-monotonic downregulation in mean expression here, which might require further examination in the context of BC subtypes^93,94^. DUSP1 (dual specificity phosphatase 1 or MAPK phosphatase 1) is a tumor-suppressor in the MAPK pathway that mediates the dephosphorylation of ERK1/2^95^, and its downregulation seen here is likely to underpin sustained proliferative signalling. FREM1 has been identified as a tumor-suppressor, whose downregulation enabled metabolic shift and tumor infiltration^96^, a finding underlined by the monotonic downregulation seen here. HFM1, helicase for meiosis 1, was reported to be altered in tumors relative to control samples^97^, and seen to be a tumor-suppressor here. ABCA10 is a member of the active transmembrane transport family, and was recently implicated in the progression-free survival of epithelial ovarian sarcoma^98^, and appears to portray a tumor-suppressor role in the context of our findings. KLK5, a serine protease, is a known tumor-suppressor whose activation is a promising anticancer therapy via repression of the mevalonate pathway^99^. The downregulation of KCNA1 (a voltage-gated potassium channel subfamily member) has been correlated with breast cancer aggressiveness^100^, lending its stage-IV salience in our analysis. KRT15 is known as cytokeratin and has recently been shown to be closely associated with tumorigenesis. Overexpression of KRT15 (cytokeratin) was seen in colorectal and squamous cell skin cancers, but its low expression in BC (as seen here) has been significantly associated with poor prognosis^101^. The remaining stage-IV salient genes were found to be involved in tumor progression via processes such as including inflammation, angiogenesis, and EMT transition.

**Figure 9:**
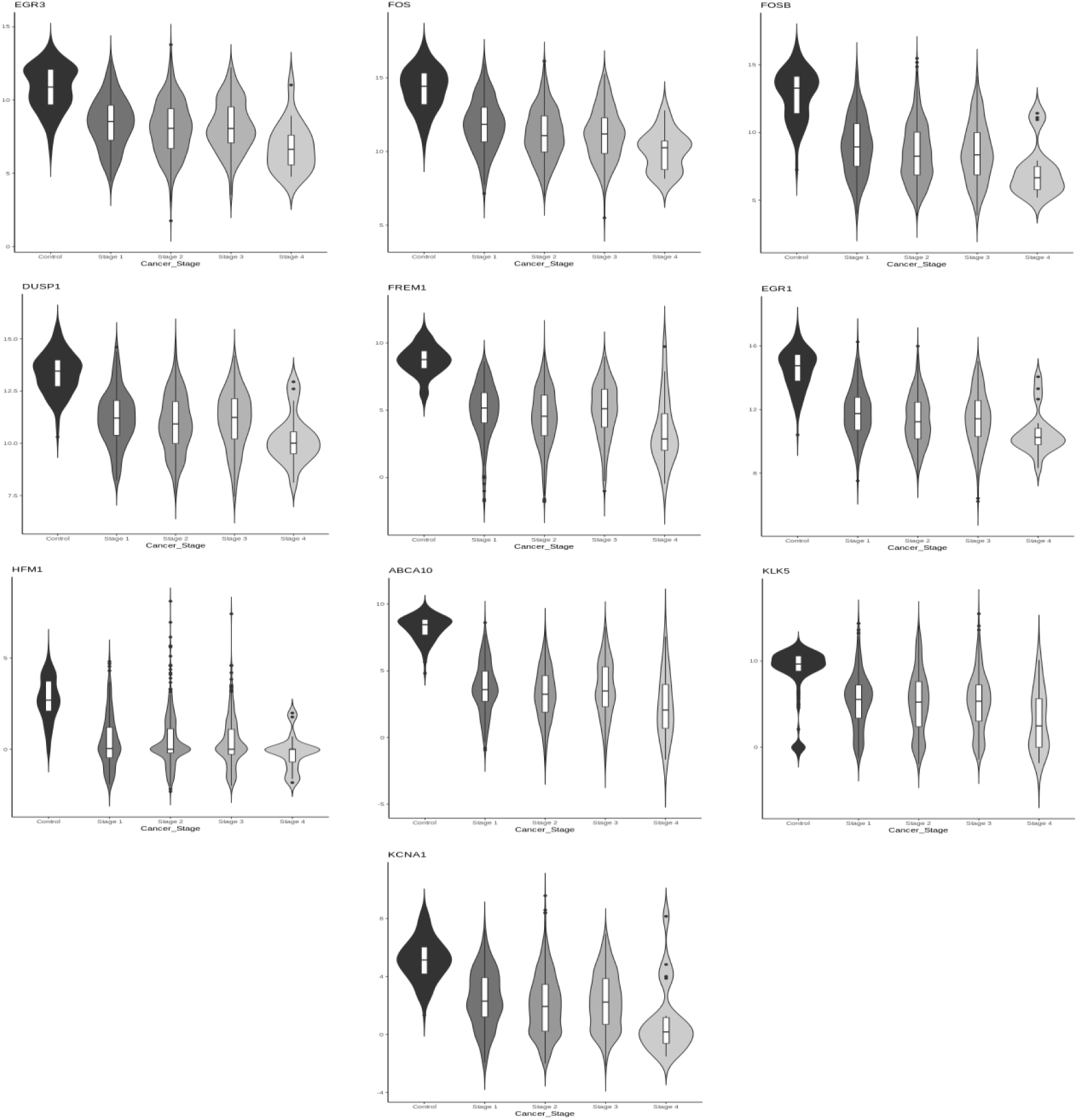
Distribution of expression patterns of the top stage-IV salient genes. The mean expression of all the genes shows monotonic downregulation, achieving an extremum in stage-IV.

The distinction between IDC and ILC has frustrated learning so far. An XGBoost model with 147 clinical, histopathological, mammogram features, and sonographic features has been reported with an internal testset accuracy of 0.84 on the binary classification problem^102^. An AutoML deep-learning approach for identifying IDC samples alone from whole slide images yielded 0.85 accuracy on an independent dataset^103^. Another study for classifying IDCs as early-stage vs late-stage yielded an AUROC of 0.47 on the external validation^104^. In this context, the external validation of our model yields a significant improvement on the state-of-the-art. However the limited sensitivity to ILC samples (conversely, specificity to IDC samples) in the external dataset presents an outstanding challenge in the histological classification of breast cancer from molecular information. Some noteworthy biomarkers from this model include: (i) CDH1 (E-cadherin), whose germline mutations were strongly associated with lobular carcinoma^105^, was found to have a specific downregulated expression signature in ILC samples; (ii) CCL14, which is known to promote angiogenesis and metastasis in breast cancer^106^, was found oncogenic in expression across both histological subtypes. To enable benchmarking against our model for this challenging problem, we have provided the code and model files for IDC vs ILC as a software repository under GPLv3 license for non-commercial purposes only (www.github.com/apalania/BC-Predict_Histological).

Commercial genomic assays for prognosticating breast-cancer adjuvant chemotherapy include Prosigna, OncotypeDX, EndoPredict (EPclin score), MammaPrint, Breast Cancer Index (exploring benefit of extension of adjuvant hormonal therapy beyond 5 years based on a 11-gene signature), HER2DX (exploring benefit of neoadjuvant systemic therapy in HER2+ BC based on a 4-gene signature)^107^, and Guardant 360^108^ and Foundation One Test^109^ (using liquid biopsies of circulating cell-free tumor DNA to profile 70+ biomarkers at progression). Scanning the signatures in these genomic assays against the ten features used in our ‘normal’ vs ‘cancer’ model yielded: two genes in common with Prosigna (FOXA1, MMP11), one gene with OncotypeDX (MMP11), one gene with HER2DX (NEK2), and one gene with Breast Cancer Index (NEK2). Scanning these signatures against the 16 features used in our molecular subtyping model yielded: four genes with Prosigna (ERBB2, FOXA1, GRB7, MLPH), four genes with HER2DX (ERBB2, GRB7, STARD3, AGR3), two with OncotypeDX (GRB7 and ERBB2), and two with Guardant360 (ERBB2, GATA3). Scanning these signatures against the 24 features used for histological subtyping yielded: one gene with Guardant360 (CDH1). Scanning these signatures against the five features used in the non-metastatic vs metastatic model did not identify anything in common. A complete discussion of the above validation is provided in Supplementary file S17.

### BC-Predict

To transition the results obtained from our studies, we developed BC-Predict which serves the models developed in a cascade inference engine and provides a comprehensive characterization of the given sample (Fig.2). The BC-predict web-server is built on Rshiny^110^ and deployed for academic research at https://apalania.shinyapps.io/BC-Predict. All predictions are accompanied by prediction probabilities to provide confidence for the predicted class. Documentation and video tutorial for the use of BC-Predict are also provided. BC-Predict generates a unified readout that could nominally support medical decision-making contingent to clinical validation and further refinement. An alternative modeling process that used a nested stratification structure instead of sequential stratification was also investigated. However this strategy did not yield an improved performance.

There are limitations and scope for improvement to our study, chiefly:

1. The metastatic model does not distinguish among the stages in pre-metastatic cancer. A refinement may be necessary to discriminate between the early-stage cancers and stage-III cancers among the pre-metastatic cancers.
2. The molecular subtype model lumps ‘Luminal A’ and ‘Luminal B’ into the ‘Luminal’ class, but these subclasses represent distinct subtypes with different prognosis and treatment.
3. The histological subtype model may be equally optimized for detecting the ILC class.
4. In addition, all models may need to be fine-tuned for distribution shifts possible in different populations, though the identity of the biomarkers could be likely invariant.

Accumulation of data is one pathway to remediate the above limitations. Initiatives like the Indian Cancer Genome Association^111^ might facilitate monitoring the performance of models such as ours on new populations.

In summary, we have developed *de novo* models to characterise breast cancer heterogeneity agnostic of hypothesis. The candidate stage-salient biomarkers could play a role in the progression of breast cancer, whose varying manifestations underlie differential response to treatment regimens. Developing models from minimal feature spaces has several advantages, chief among them being sensitivity to heterogeneous individual presentation, and generalization to out-of-domain population. One example of this in the present study is the performant external validation of the Molecular Subtype model on the TNBC-only African-enriched multiethnic international cohort (25/26 samples correctly identified). It is remarkable that TNBC is also the most common molecular subtype in the Indian subcontinent. It also frustrates drug discovery programs with few druggable targets. The candidate biomarkers identified here could provide novel hypotheses for chemotherapy and immunotherapy investigations. Our study overcomes certain limitations of earlier models, namely reporting of balanced performance metrics, availability for academic research, and external validation. The confidence returned by BC-Predict predictions could be used to safeguard against weak or uncertain evidence, necessary to combat the hazard with AI/ML modelling^112^. The clinical translation of AI/ML models would be a step forward for personalized medicine, necessitating adequate regulation to ensure the benefits of AI for all^113,114^. Validation and assurance of model quality alleviate the risks of distribution drift and cohort selection bias, and pave the way for clinically effective decision support aids in precision oncology centers. The realisation of software-as-medical-devices promises to revolutionize the diagnosis, triage, and treatment of cancers.

### Conclusions

Assessment of low-risk genetic factors unmasks induced vulnerabilities, and early-stage characterization of breast cancer heterogeneity constitutes the premise for personalized and targeted precision medicine. In this work, we have developed *de novo* models for addressing key problems in breast cancer heterogeneity based on public-domain expression datasets. Using custom protocols to identify biomarkers of interest to each problem, we have trained, optimised and externally validated the models. Our analysis has yielded novel and stage-salient drivers of cancer progression, including two stage-I salient genes (CHRNA6, MMP10), two stage-II salient genes (DEPDC1, COXA1), ten stage-III salient genes (including AKR7A3, FOXA1, CXCL5 and GDF5) and 20 stage-IV salient genes (including FREM1 and HFM1). We have developed solutions to four problems of interest in characterizing breast cancer heterogeneity: (i) ‘cancer’ vs ‘normal’ based on 10 features (2 stage salient genes and 8 top linear model genes) with balanced accuracy ∼ 97.42% on external validation; (ii) non-metastatic vs metastatic based on 5 features with balanced accuracy ∼ 88.22% on external validation; (iii) molecular subtyping (namely Luminal, HER2+, and TNBC) based on 16 features with balanced accuracy ∼88.79% on external validation; and (iv) histological subtyping (IDC vs ILC) based on 24 features with ensemble accuracy ∼94.23% on external validation. We have validated our results in multiple modalities. Based on these outcomes, we have developed an inference engine BC-Predict, which serves the best models developed for each problem, upon an input instance of expression data from a patient sample. BC-Predict is available for academic and non-commercial purposes as an experimental predictive aid for characterization of breast cancer heterogeneity based on minimal expression information, and subject to refinement with new knowledge. In conclusion, we have identified various novel candidate biomarkers of heterogeneous breast cancers that have been embedded into an integrated and validated cascade model towards the ultimate aim of expediting personalized differential diagnosis and cure.

## Data Availability

All the Supplementary Information for this study are available at: https://doi.org/10.6084/m9.figshare.25282906. BC-Predict is available for academic research at: https://apalania.shinyapps.io/BC-Predict. Source and object code of the ensemble classifier for histological subtyping (BC-Predict_Histological) is available at: https://github.com/apalania/BC-Predict_Histological for non-commercial uses only. A demo of BC-Predict is available at: https://www.youtube.com/watch?v=NDr39vXWS6U.

## Acknowledgements

We are grateful to Dr Natarajan Vaithilingam MS (Gen Surg); FRCSEd; FEBS (Br Surg European Board), Consultant Surgeon NHS–UK, for helpful discussions and comments. We would like to thank the Management of SASTRA Deemed University for infrastructure and support. This study makes use of the TCGA dataset (generated by The Cancer Genome Atlas Consortium), METABRIC dataset (generated by the Molecular Taxonomy of Breast Cancer International Consortium), ICGC dataset (generated by International Cancer Genome Consortium), and GEO datasets. This work was supported in part on DST-SERB grant EMR/2017/000470. Computing in our lab is also supported on a Google TPU Research Cloud (TRC) grant of Cloud TPU VMs.

## References

1 Siegel RL, Giaquinto AN, Jemal A. Cancer statistics, 2024. CA Cancer J Clin. 2024; 74(1):12–49. doi: 10.3322/caac.21820.

2 Sung H, Ferlay J, Siegel RL, et al. Global Cancer Statistics 2020: GLOBOCAN Estimates of Incidence and Mortality Worldwide for 36 Cancers in 185 Countries. CA Cancer J Clin. 2021 May;71(3):209–249. doi: 10.3322/caac.21660.

3 Cassidy J, Bissett D, Payne M. and Morris-Stiff, G. eds., 2015. Oxford handbook of oncology. OUP Oxford.

4 Hanahan D. Hallmarks of Cancer: New Dimensions. Cancer Discov. 2022; 12(1):31–46. doi: 10.1158/2159-8290.CD-21-1059.

5 Risch HA, McLaughlin JR, Cole DE, et al. Population BRCA1 and BRCA2 Mutation Frequencies and Cancer Penetrances: A Kin–Cohort Study in Ontario, Canada, JNCI: Journal of the National Cancer Institute. 2006; 98(23):1694–1706. doi: 10.1093/jnci/djj465

6 Allain DC. Genetic counseling and testing for common hereditary breast cancer syndromes: a paper from the 2007 William Beaumont hospital symposium on molecular pathology. J Mol Diagn. 2008; 10(5):383–95. doi: 10.2353/jmoldx.2008.070161.

7 Lindor NM, McMaster ML, Lindor CJ, Greene MH; National Cancer Institute, Division of Cancer Prevention, Community Oncology and Prevention Trials Research Group. Concise handbook of familial cancer susceptibility syndromes – second edition. J Natl Cancer Inst Monogr. 2008; (38):1–93. doi: 10.1093/jncimonographs/lgn001.

8 Manyonda I, Sinai Talaulikar V, Pirhadi R, Ward J, Banerjee D, Onwude J. Could Perimenopausal Estrogen Prevent Breast Cancer? Exploring the Differential Effects of Estrogen-Only Versus Combined Hormone Replacement Therapy. J Clin Med Res. 2022; 14(1):1–7. doi: 10.14740/jocmr4646.

9 Bhattacharyya GS, Doval CD, Desai CJ, et al. Overview of Breast Cancer and Implications of Overtreatment of Early-Stage Breast Cancer: An Indian Perspective. JCO global oncology. 2020; 6:789–798. doi: 10.1200/GO.20.00033.

10 Malvia S, Bagadi SA, Dubey US, Saxena S. Epidemiology of breast cancer in Indian women. Asia-Pacific journal of clinical oncology. 2017; 13(4):289–295. doi: 10.1111/ajco.12661.

11 McKinney SM, Sieniek M, Godbole V et al. International evaluation of an AI system for breast cancer screening. Nature. 2020; 577(7788):89–94.

12 Mostavi M, Chiu YC, Huang Y, Chen Y. Convolutional neural network models for cancer type prediction based on gene expression. BMC Med Genomics. 2020; 13(Suppl 5):44. doi: 10.1186/s12920-020-0677-2.

13 Zhao Y, Pan Z, Namburi S et al. CUP-AI-Dx: A tool for inferring cancer tissue of origin and molecular subtype using RNA gene-expression data and artificial intelligence. EBioMedicine. 2020; 61:103030. doi: 10.1016/j.ebiom.2020.103030.

14 Bastien RR, Rodríguez-Lescure Á, Ebbert MT et al. PAM50 breast cancer subtyping by RT-qPCR and concordance with standard clinical molecular markers. BMC Med Genomics. 2012; 5:44. doi: 10.1186/1755-8794-5-44.

15 Gao F, Wang W, Tan, M. et al. DeepCC: a novel deep learning-based framework for cancer molecular subtype classification. Oncogenesis. 2019; 8:44. doi: 10.1038/s41389-019-0157-8.

16 Mohaiminul Islam M, Huang S, Ajwad R, Chi C, Wang Y, Hu P. An integrative deep learning framework for classifying molecular subtypes of breast cancer. Comput Struct Biotechnol J. 2020; 18:2185–2199. doi: 10.1016/j.csbj.2020.08.005.

17 Zhang S, Fitzsimmons KC, Hurvitz SA. Oncotype DX Recurrence Score in premenopausal women. Ther Adv Med Oncol. 2022; 14:17588359221081077. doi: 10.1177/17588359221081077.

18 Almstedt K, Mendoza S, Otto M, et al. EndoPredict® in early hormone receptor-positive, HER2-negative breast cancer. Breast Cancer Res Treat. 2020; 182(1):137–146. doi: 10.1007/s10549-020-05688-1.

19 Soliman, H., Shah, V., Srkalovic, G. et al. MammaPrint guides treatment decisions in breast Cancer: results of the IMPACt trial. BMC Cancer. 2020;20:81. doi: 10.1186/s12885-020-6534-z.

20 Baskota SU, Dabbs DJ, Clark BZ, Bhargava R. Prosigna® breast cancer assay: histopathologic correlation, development, and assessment of size, nodal status, Ki-67 (SiNK™) index for breast cancer prognosis. Mod Pathol. 2021; 34(1):70–76. doi: 10.1038/s41379-020-0643-8.

21 Bartlett JMS, Sgroi DC, Treuner K, Zhang Y, Ahmed I, Piper T, Salunga R, Brachtel EF, Pirrie SJ, Schnabel CA, Rea DW. Breast Cancer Index and prediction of benefit from extended endocrine therapy in breast cancer patients treated in the Adjuvant Tamoxifen-To Offer More? (aTTom) trial. Ann Oncol. 2019; 30(11):1776–1783. doi: 10.1093/annonc/mdz289.

22 Muthamilselvan S, Ramasami Sundhar Baabu P, Palaniappan A. Microfluidics for Profiling miRNA Biomarker Panels in AI-Assisted Cancer Diagnosis and Prognosis. Technology in Cancer Research & Treatment. 2023; 22. doi: 10.1177/15330338231185284.

23 Guler EN. Gene Expression Profiling in Breast Cancer and Its Effect on Therapy Selection in Early-Stage Breast Cancer. European journal of breast health. 2017; 13(4):168–174. doi: 10.5152/ejbh.2017.3636.

24 Brierley J, Gospodarowicz M, O’Sullivan B. The principles of cancer staging. Ecancermedicalscience. 2016; 10:ed61. doi: 10.3332/ecancer.2016.ed61.

25 Kourou, K., Exarchos, T. P., Exarchos, K. P., Karamouzis, M. V. & Fotiadis, D. I. Machine learning applications in cancer prognosis and prediction. Computational and structural biotechnology journal. 2014; 13:8–17. doi: 10.1016/j.csbj.2014.11.005.

26 Horr C, Buechler SA. Breast Cancer Consensus Subtypes: A system for subtyping breast cancer tumors based on gene expression. NPJ Breast Cancer. 2021; 7(1):136. doi: 10.1038/s41523-021-00345-2.

27 Johnson KS, Conant EF, Soo MS. Molecular Subtypes of Breast Cancer: A Review for Breast Radiologists. Journal of Breast Imaging. 2021; 3(1):12–24. doi: 10.1093/jbi/wbaa110.

28 Vaidya JS, Massarut S, Vaidya HJ, Alexander EC, Richards T, Caris JA, Sirohi B, Tobias JS. Rethinking neoadjuvant chemotherapy for breast cancer. BMJ. 2018; 360:j5913. doi: 10.1136/bmj.j5913.

29 Fu D, Zuo Q, Huang Q et al. Molecular Classification of Lobular Carcinoma of the Breast. Sci Rep. 2017;7:43265. doi: 10.1038/srep43265.

30 Fitzgibbons PL, Page DL, Weaver D et al. Prognostic factors in breast cancer. College of American Pathologists Consensus Statement 1999. Arch Pathol Lab Med. 2000; 124(7):966–78. doi: 10.5858/2000-124-0966-PFIBC.

31 Rakha EA, Reis-Filho JS, Baehner F et al. Breast cancer prognostic classification in the molecular era: the role of histological grade. Breast Cancer Res. 2010; 12(4):207. doi: 10.1186/bcr2607.

32 Sarathi, A. & Palaniappan, A. Novel significant stage-specific differentially expressed genes in hepatocellular carcinoma. BMC Cancer. 2019; 19:663. doi: 10.1186/s12885-019-5838-3.

33 Broad Institute TCGA Genome Data Analysis Center. Analysis-ready standardized TCGA data from broad GDAC firehose 2016_01_28 run. Broad institute of MIT and Harvard. Dataset; 2016. doi: 10.7908/C11G0KM9.

34 Giuliano AE, Edge SB, Hortobagyi GN. Eighth Edition of the AJCC Cancer Staging Manual: Breast Cancer. Ann Surg Oncol. 2018; 25(7):1783–1785. doi: 10.1245/s10434-018-6486-6.

35 Dai, Xiaofeng et al. Breast cancer intrinsic subtype classification, clinical use and future trends. American journal of cancer research. 2015; 5(10): 2929–43.

36 Weigelt B, Geyer FC, Reis-Filho JS. Histological types of breast cancer: how special are they? Mol Oncol. 2010; 4(3):192–208. doi: 10.1016/j.molonc.2010.04.004.

37 Winchester DJ, Chang HR, Graves TA, et al. A comparative analysis of lobular and ductal carcinoma of the breast: presentation, treatment, and outcomes. J Am Coll Surg. 1998; 186(4):416–22. doi: 10.1016/s1072-7515(98)00051-9.

38 Law CW, Chen Y, Shi W, Smyth GK. voom: Precision weights unlock linear model analysis tools for RNA-seq read counts. Genome Biol. 2014;15(2):R29. doi: 10.1186/gb-2014-15-2-r29.

39 Nitesh V. Chawla, Kevin W. Bowyer, Lawrence O. Hall, and W. Philip Kegelmeyer. SMOTE: synthetic minority over-sampling technique. J. Artif. Int. Res. 2002; 16(1):321–357.

40 Ritchie ME, Phipson B, Wu D, et al. limma powers differential expression analyses for RNA-sequencing and microarray studies. Nucleic Acids Research. 2015; 43:e47–e47, doi:10.1093/nar/gkv007.

41 Kursa MB, Rudnicki WR. Feature Selection with the Boruta Package. Journal of Statistical Software. 2010; 36(11):1–13. http://www.jstatsoft.org/v36/i11/.

42. International Cancer Genome Consortium, Hudson TJ, Anderson W, et al. International network of cancer genome projects. Nature. 2010; 464(7291):993–998. doi: 10.1038/nature08987.

43. The Genotype-Tissue Expression (GTEx) Project was supported by the Common Fund of the Office of the Director of the National Institutes of Health, and by NCI, NHGRI, NHLBI, NIDA, NIMH, and NINDS. The data used for the analyses described in this were obtained from: GTEx_Analysis_2017-06-05_v8_RNASeQCv1.1.9_gene_tpm.gct.gz the GTEx Portal and/or dbGaP accession number phs000424.v8.p2.

44 Barrett T, Wilhite SE, Ledoux P, et al. A.NCBI GEO: archive for functional genomics data sets--update. Nucleic Acids Res. 2013; 41(Database issue):D991–5.

45 Curtis C, Shah SP, Chin SF, et al. METABRIC Group; The genomic and transcriptomic architecture of 2,000 breast tumours reveals novel subgroups. Nature. 2012; 486(7403):346–52. doi: 10.1038/nature10983.

46 Franks JM, Cai G, Whitfield ML. Feature specific quantile normalization enables cross-platform classification of molecular subtypes using gene expression data. Bioinformatics. 2018; 34(11):1868–1874. doi: 10.1093/bioinformatics/bty026.

47 Martini R, Delpe P, Chu TR, et al. African Ancestry-Associated Gene Expression Profiles in Triple-Negative Breast Cancer Underlie Altered Tumor Biology and Clinical Outcome in Women of African Descent. Cancer Discov. 2022; 12(11):2530–2551. doi: 10.1158/2159-8290.CD-22-0138.

48 The Metastatic Breast Cancer Project (Provisional, February 2020). The cBioPortal for Cancer Genomics https://identifiers.org/cbioportal:brca_mbcproject_wagle_2017 (2020)

49 Ru Y, Kechris KJ, Tabakoff B, Hoffman P, Radcliffe RA, Bowler R, Mahaffey S, Rossi S, Calin GA, Bemis L, Theodorescu D. The multiMiR R package and database: integration of microRNA-target interactions along with their disease and drug associations. Nucleic Acids Res. 2014;42(17):e133. doi: 10.1093/nar/gku631.

50 Agarwal V, Bell GW, Nam JW, Bartel DP. Predicting effective microRNA target sites in mammalian mRNAs. Elife. 2015 Aug 12;4:e05005. doi: 10.7554/eLife.05005.

51 Wang X. miRDB: a microRNA target prediction and functional annotation database with a wiki interface. RNA. 2008 Jun;14(6):1012–7. doi: 10.1261/rna.965408.

52. 52 Enright, A.J., John, B., Gaul, U., et al. MicroRNA targets in Drosophila. Genome Biol 5, R1 (2003). 10.1186/gb-2003-5-1-r1

53 Huang HY, Lin YC, Cui S, Huang Y, Tang Y, Xu J, Bao J, Li Y, Wen J, Zuo H, Wang W, Li J, Ni J, Ruan Y, Li L, Chen Y, Xie Y, Zhu Z, Cai X, Chen X, Yao L, Chen Y, Luo Y, LuXu S, Luo M, Chiu CM, Ma K, Zhu L, Cheng GJ, Bai C, Chiang YC, Wang L, Wei F, Lee TY, Huang HD. miRTarBase update 2022: an informative resource for experimentally validated miRNA-target interactions. Nucleic Acids Res. 2022; 50(D1):D222–D230. doi: 10.1093/nar/gkab1079.

54 Cedoz PL, Prunello M, Brennan K, Gevaert O. MethylMix 2.0: an R package for identifying DNA methylation genes. Bioinformatics. 2018; 34(17):3044–3046. doi: 10.1093/bioinformatics/bty156.

55 Muthamilselvan S, Raghavendran A, Palaniappan A (2022) Stage-differentiated ensemble modeling of DNA methylation landscapes uncovers salient biomarkers and prognostic signatures in colorectal cancer progression. PLoS ONE 17(2): e0249151. doi: 10.1371/journal.pone.0249151.

56 Lex A, Gehlenborg N, Strobelt H, Vuillemot R, Pfister H. UpSet: Visualization of Intersecting Sets. IEEE Trans Vis Comput Graph. 2014; 20(12):1983–92. doi: 10.1109/TVCG.2014.2346248.

57 Thissen, D., Steinberg, L. & Kuang, D. Quick and easy implementation of the Benjamini-Hochberg procedure for controlling the false positive rate in multiple comparisons. Journal of educational and behavioral statistics. 2002;27:77–83.

58 Muthamilselvan S, Palaniappan A. BrcaDx: precise identification of breast cancer from expression data using a minimal set of features. Front Bioinform. 2023;3:1103493. doi: 10.3389/fbinf.2023.1103493.

59 Taghizadeh E, Heydarheydari S, Saberi A, JafarpoorNesheli S, Rezaeijo SM. Breast cancer prediction with transcriptome profiling using feature selection and machine learning methods. BMC Bioinformatics. 2022; 23(1):410. doi: 10.1186/s12859-022-04965-8.

60 Jin W, Chen BB, Li JY, et al. TIEG1 inhibits breast cancer invasion and metastasis by inhibition of epidermal growth factor receptor (EGFR) transcription and the EGFR signaling pathway. Mol Cell Biol. 2012; 32(1):50–63. doi: 10.1128/MCB.06152-11.

61 Subramaniam M, Hefferan TE, Tau K, et al. Tissue, cell type, and breast cancer stage-specific expression of a TGF-beta inducible early transcription factor gene. J Cell Biochem. 1998;68(2):226–36.

62 Wu K, Weng Z, Tao Q, et al. Stage-specific expression of breast cancer-specific gene gamma-synuclein. Cancer Epidemiol Biomarkers Prev. 2003; 12(9):920–5.

63 Croteau DL, Singh DK, Hoh Ferrarelli L, Lu H, Bohr VA. RECQL4 in genomic instability and aging. Trends Genet. 2012; 28(12):624–31. doi: 10.1016/j.tig.2012.08.003.

64 Luong TT, Li Z, Priedigkeit N, et al. Hrq1/RECQL4 regulation is critical for preventing aberrant recombination during DNA intrastrand crosslink repair and is upregulated in breast cancer. PLoS Genet. 2022; 18(9):e1010122. doi: 10.1371/journal.pgen.1010122.

65 Arora A, Agarwal D, Abdel-Fatah TM, et al. RECQL4 helicase has oncogenic potential in sporadic breast cancers. J Pathol. 2016; 238(4):495–501. doi: 10.1002/path.4681.

66 Masuda H, Zhang D, Bartholomeusz C, et al. Role of epidermal growth factor receptor in breast cancer. Breast Cancer Res Treat. 2012; 136(2):331–45. doi: 10.1007/s10549-012-2289-9.

67 Ali R, Wendt MK. The paradoxical functions of EGFR during breast cancer progression. Signal Transduct Target Ther. 2017; 2:16042. doi: 10.1038/sigtrans.2016.42.

68 Cao Y, Chu C, Li X, Gu S, Zou Q, Jin Y. RNA-binding protein QKI suppresses breast cancer via RASA1/MAPK signaling pathway. Ann Transl Med. 2021; 9(2):104. doi: 10.21037/atm-20-4859

69 Gu S, Chu C, Chen W, et al. Prognostic value of epithelial-mesenchymal transition related genes: SLUG and QKI in breast cancer patients. Int J Clin Exp Pathol. 2019; 12(6):2009–2021.

70 Wei Y, Wang X, Zhang Z, Xie M, Li Y, Cao H, Zhao X. Role of Polymorphisms of FAM13A, PHLDB1, and CYP24A1 in Breast Cancer Risk. Curr Mol Med. 2019;19(8):579–588. doi: 10.2174/1566524019666190619125109

71 Liu T, Fang Y. Research for Expression and Prognostic Value of GABRD in Colon Cancer and Coexpressed Gene Network Construction Based on Data Mining. Comput Math Methods Med. 2021; 2021:5544182. doi: 10.1155/2021/5544182.

72 Song L, Chang R, Dai C, et al. SORBS1 suppresses tumor metastasis and improves the sensitivity of cancer to chemotherapy drug. Oncotarget. 2017;8(6):9108–9122. doi: 10.18632/oncotarget.12851.

73 Uhlen M, Zhang C, Lee S, et al. A pathology atlas of the human cancer transcriptome. Science. 2017; 357(6352):eaan2507. doi: 10.1126/science.aan2507.

74 Singh S, Pillai S, Chellappan S. Nicotinic acetylcholine receptor signaling in tumor growth and metastasis. J Oncol. 2011; 2011:456743. doi: 10.1155/2011/456743.

75 Lam DC, Girard L, Ramirez R, et al. Expression of nicotinic acetylcholine receptor subunit genes in non-small-cell lung cancer reveals differences between smokers and nonsmokers. Cancer Res. 2007; 67(10):4638–47. doi: 10.1158/0008-5472.CAN-06-4628.

76 Zhang G, Miyake M, Lawton A, Goodison S, Rosser CJ. Matrix metalloproteinase-10 promotes tumor progression through regulation of angiogenic and apoptotic pathways in cervical tumors. BMC Cancer. 2014; 14:310. doi: 10.1186/1471-2407-14-310.

77 Piskór BM, Przylipiak A, Dąbrowska E, et al. Plasma Level of MMP-10 May Be a Prognostic Marker in Early Stages of Breast Cancer. J Clin Med. 2020; 9(12):4122. doi: 10.3390/jcm9124122.

78 Mi Y, Zhang C, Bu Y, et al. DEPDC1 is a novel cell cycle related gene that regulates mitotic progression. BMB Rep. 2015; 48(7):413–8. doi: 10.5483/bmbrep.2015.48.7.036.

79 Zhao H, Yu M, Sui L, et al. High Expression of DEPDC1 Promotes Malignant Phenotypes of Breast Cancer Cells and Predicts Poor Prognosis in Patients With Breast Cancer. Front Oncol. 2019; 9:262. doi: 10.3389/fonc.2019.00262.

80 Zhang L, Du Y, Xu S, et al. DEPDC1, negatively regulated by miR-26b, facilitates cell proliferation via the up-regulation of FOXM1 expression in TNBC. Cancer Lett. 2019; 442:242–251. doi: 10.1016/j.canlet.2018.11.003.

81 He Z, Wang F, Zhang W, Ding J, Ni S. Comprehensive and integrative analysis identifies COX7A1 as a critical methylation-driven gene in breast invasive carcinoma. Ann Transl Med. 2019; 7(22):682. doi: 10.21037/atm.2019.11.97.

82 Williams S, Bateman A, O’Kelly I. Altered expression of two-pore domain potassium (K2P) channels in cancer. PLoS One. 2013; 8(10):e74589. doi: 10.1371/journal.pone.0074589.

83 Dookeran KA, Zhang W, Stayner L, Argos M. Associations of two-pore domain potassium channels and triple negative breast cancer subtype in The Cancer Genome Atlas: systematic evaluation of gene expression and methylation. BMC Res Notes. 2017; 10(1):475. doi: 10.1186/s13104-017-2777-4.

84 Kanda M, Shimizu D, Tanaka H, et al. Metastatic pathway-specific transcriptome analysis identifies MFSD4 as a putative tumor suppressor and biomarker for hepatic metastasis in patients with gastric cancer. Oncotarget. 2016; 7(12):13667–79. doi: 10.18632/oncotarget.7269.

85 Tervasmäki A, Winqvist R, Jukkola-Vuorinen A, Pylkäs K. Recurrent CYP2C19 deletion allele is associated with triple-negative breast cancer. BMC Cancer. 2014; 14:902. doi: 10.1186/1471-2407-14-902.

86 Romero-Moreno, R., Curtis, K.J., Coughlin, T.R. et al. The CXCL5/CXCR2 axis is sufficient to promote breast cancer colonization during bone metastasis. Nat Commun. 2019; 10:4404. doi: 10.1038/s41467-019-12108-6.

87 Hlaváč V, Brynychová V, Václavíková R, et al. The role of cytochromes p450 and aldo-keto reductases in prognosis of breast carcinoma patients. Medicine (Baltimore). 2014; 93(28):e255. doi: 10.1097/MD.0000000000000255.

88 Makoukji J, Raad M, Genadry K, et al. Association between CLN3 (Neuronal Ceroid Lipofuscinosis, CLN3 Type) Gene Expression and Clinical Characteristics of Breast Cancer Patients. Front Oncol. 2015; 5:215. doi: 10.3389/fonc.2015.00215.

89 Margheri F, Schiavone N, Papucci L, et al. GDF5 regulates TGFß-dependent angiogenesis in breast carcinoma MCF-7 cells: in vitro and in vivo control by anti-TGFß peptides. PLoS One. 2012; 7(11):e50342. doi: 10.1371/journal.pone.0050342.

90 Fu X, Pereira R, De Angelis C, et al. FOXA1 upregulation promotes enhancer and transcriptional reprogramming in endocrine-resistant breast cancer. Proc Natl Acad Sci U S A. 2019; 116(52):26823–26834. doi: 10.1073/pnas.1911584116.

91 Blanco MA, LeRoy G, Khan Z, et al. Global secretome analysis identifies novel mediators of bone metastasis. Cell Res. 2012; 22(9):1339–55. doi: 10.1038/cr.2012.89.

92 Suzuki T, Inoue A, Miki Y, et al. Early growth responsive gene 3 in human breast carcinoma: a regulator of estrogen-meditated invasion and a potent prognostic factor. Endocr Relat Cancer. 2007; 14(2):279–92. doi: 10.1677/ERC-06-0005.

93 Lu C, Shen Q, DuPré E, Kim H, Hilsenbeck S, Brown PH. cFos is critical for MCF-7 breast cancer cell growth. Oncogene. 2005; 24(43):6516–24. doi: 10.1038/sj.onc.1208905.

94 Bamberger AM, Methner C, Lisboa BW, Städtler C, Schulte HM, Löning T, Milde-Langosch K. Expression pattern of the AP-1 family in breast cancer: association of fosB expression with a well-differentiated, receptor-positive tumor phenotype. Int J Cancer. 1999; 84(5):533–8. doi: 10.1002/(sici)1097-0215(19991022)84:5<533::aid-ijc16>3.0.co;2-j.

95 Chen CC, Hardy DB, Mendelson CR. Progesterone receptor inhibits proliferation of human breast cancer cells via induction of MAPK phosphatase 1 (MKP-1/DUSP1). J Biol Chem. 2011; 286(50):43091–102. doi: 10.1074/jbc.M111.295865.

96 Li HN, Li XR, Lv ZT, Cai MM, Wang G, Yang ZF. Elevated expression of FREM1 in breast cancer indicates favorable prognosis and high-level immune infiltration status. Cancer Med. 2020; 9(24):9554–9570. doi: 10.1002/cam4.3543.

97 Taylor BS, Barretina J, Socci ND, et al. Functional copy-number alterations in cancer. PLoS One. 2008; 3(9):e3179. doi: 10.1371/journal.pone.0003179.

98 Seborova K, Vaclavikova R, Soucek P, et al. Association of ABC gene profiles with time to progression and resistance in ovarian cancer revealed by bioinformatics analyses. Cancer Med. 2019; 8(2):606–616. doi: 10.1002/cam4.1964.

99 Pampalakis G, Obasuyi O, Papadodima O, Chatziioannou A, Zoumpourlis V, Sotiropoulou G. The KLK5 protease suppresses breast cancer by repressing the mevalonate pathway. Oncotarget. 2014; 5(9):2390–403. doi: 10.18632/oncotarget.1235.

100 Lallet-Daher H, Wiel C, Gitenay D, et al. Potassium channel KCNA1 modulates oncogene-induced senescence and transformation. Cancer Res. 2013; 73(16):5253–65. doi: 10.1158/0008-5472.CAN-12-3690.

101 Zhong P, Shu R, Wu H, Liu Z, Shen X, Hu Y. Low KRT15 expression is associated with poor prognosis in patients with breast invasive carcinoma. Exp Ther Med. 2021; 21(4):305. doi: 10.3892/etm.2021.9736.

102 Vy VPT, Yao MM-S, Khanh Le NQ, Chan WP. Machine Learning Algorithm for Distinguishing Ductal Carcinoma In Situ from Invasive Breast Cancer. Cancers. 2022; 14(10):2437. doi: 10.3390/cancers14102437.

103 Zeng Y, Zhang J. A machine learning model for detecting invasive ductal carcinoma with Google Cloud AutoML Vision. Computers in Biology and Medicine. 2020;122: 103861.doi: 10.1016/j.compbiomed.2020.103861.

104 Roy S, Kumar R, Mittal V, Gupta D. Classification models for Invasive Ductal Carcinoma Progression, based on gene expression data-trained supervised machine learning. Sci Rep. 2020; 10(1):4113. doi: 10.1038/s41598-020-60740-w.

105 Corso G, Veronesi P, Sacchini V, Galimberti V. Prognosis and outcome in CDH1-mutant lobular breast cancer. Eur J Cancer Prev. 2018; 27(3):237–238. doi: 10.1097/CEJ.0000000000000405.

106 Li Q, Shi L, Gui B, et al. Binding of the JmjC demethylase JARID1B to LSD1/NuRD suppresses angiogenesis and metastasis in breast cancer cells by repressing chemokine CCL14. Cancer Res. 2011; 71(21):6899–908. doi: 10.1158/0008-5472.CAN-11-1523.

107 Prat A, Guarneri V, Pascual T, et al. Development and validation of the new HER2DX assay for predicting pathological response and survival outcome in early-stage HER2-positive breast cancer. EBioMedicine. 2022; 75:103801. doi: 10.1016/j.ebiom.2021.103801.

108 Guardant Health Guardant360 CDx first FDA-approved liquid biopsy for comprehensive tumor mutation profiling across all solid cancers. News release. Guardant Health; August 7, 2020. Accessed February 1, 2021. https://investors.guardanthealth.com/press-releases/press-releases/2020/Guardant-Health-Guardant360-CDx-First-FDA-Approved-Liquid-Biopsy-for-Comprehensive-Tumor-Mutation-Profiling-Across-All-Solid-Cancers/default.aspx

109 FDA approves Foundation Medicine’s FoundationOne Liquid CDx, a comprehensive pan-tumor liquid biopsy test with multiple companion diagnostic indications for patients with advanced cancer. News release. Foundation Medicine; October 8, 2020. Accessed February 1, 2024. https://www.foundationmedicine.com/press-releases/fda-approves-foundation-medicine’s-foundationone%C2%AEliquid-cdx,-a-comprehensive-pan-tumor-liquid-biopsy-test-with-multiple-companion-diagnostic-indications-for-patients-with-advanced-cancer

110. 110 Chang W., Cheng J., Allaire J., Sievert C., Schloerke B., Xie Y., et al. (2023). shiny: Web Application Framework for R. R package version 1.7.4. Available at: https://shiny.rstudio.com/.

111 Dixit S, Sadanandam A. The 2nd Conference and Workshop of The Cancer Genome Atlas (TCGA) in India: Towards Team Science for Multi-omics Cancer Research in South Asia. Ecancermedicalscience. 2021; 15:ed111. doi: 10.3332/ecancer.2021.ed111.

112 Yao K, Tong CY, Cheng C. A framework to predict the applicability of Oncotype DX, MammaPrint, and E2F4 gene signatures for improving breast cancer prognostic prediction. Sci Rep. 2022; 12(1):2211. doi: 10.1038/s41598-022-06230-7.

113 El Naqa I, Karolak A, Luo Y. et al. Translation of AI into oncology clinical practice. Oncogene. 2023; 42:3089–3097. doi:10.1038/s41388-023-02826-z.

114 Hickman SE, Baxter GC, Gilbert FJ. Adoption of artificial intelligence in breast imaging: evaluation, ethical constraints and limitations. Br J Cancer. 2021; 125(1):15–22. doi: 10.1038/s41416-021-01333-w.

